# Germline predisposition to pediatric Ewing sarcoma is uniquely characterized by inherited pathogenic variants in DNA damage repair genes

**DOI:** 10.1101/2022.01.07.22268685

**Authors:** Riaz Gillani, Sabrina Y. Camp, Seunghun Han, Jill K. Jones, Schuyler O’Brien, Erin L. Young, Lucy Hayes, Gareth Mitchell, Trent Fowler, Alexander Gusev, Junne Kamihara, Katherine A. Janeway, Joshua D. Schiffman, Brian D. Crompton, Saud H. AlDubayan, Eliezer M. Van Allen

**Affiliations:** Department of Pediatric Oncology, Dana-Farber Cancer Institute, Boston, MA 02215, USA; Broad Institute of Harvard and MIT, Cambridge, MA 02142, USA; Department of Pediatrics, Harvard Medical School, Boston, MA 02215, USA; Boston Children’s Hospital, Boston, MA 02115, USA; Department of Medical Oncology, Dana-Farber Cancer Institute, Boston, MA 02115, USA; Harvard Medical School, Boston, MA 02115, USA; Huntsman Cancer Institute, 2000 Circle of Hope Dr, Room 4246, Salt Lake City, UT 84112; Department of Epidemiology, Harvard T.H. Chan School of Public Health, Boston, Massachusetts, USA; Division of Genetics, Brigham and Women’s Hospital, Boston, MA 02115; College of Medicine, King Saudi bin Abdulaziz University for Health Sciences, Riyadh, Saudi Arabia; Center for Cancer Genomics, Dana-Farber Cancer Institute, Boston, MA 02115, USA

## Abstract

More knowledge is needed around the role and importance of specific genes in germline predisposition to Ewing sarcoma to inform biological investigation and clinical practice. In this study, we evaluated the enrichment of pathogenic germline variants in Ewing sarcoma relative to other pediatric sarcoma subtypes, as well as patterns of inheritance of these variants. We carried out an ancestry-matched case-control analysis to screen for enrichment of pathogenic germline variants in 141 established cancer predisposition genes in 1138 individuals with pediatric sarcoma diagnoses (222 Ewing sarcoma cases) relative to identically processed cancer-free controls. Findings in Ewing sarcoma were validated with an additional cohort of 425 individuals, and 301 Ewing sarcoma parent-proband trios were analyzed for inheritance patterns of identified pathogenic variants. A distinct pattern of pathogenic germline variants was seen in Ewing sarcoma relative to other sarcoma subtypes. *FANCC* was the only gene with an enrichment signal for heterozygous pathogenic variants in the discovery Ewing sarcoma cohort (OR 14.4, 95% CI 3.5 – 51.2, p = 0.002, FDR = 0.28). This enrichment in *FANCC* heterozygous pathogenic variants was seen again in the Ewing sarcoma validation cohort (OR 5.1, 95% CI 1.2 – 18.5, p = 0.03, single hypothesis), representing a broader importance of genes involved in DNA damage repair, which were also nominally enriched in Ewing sarcoma cases. Pathogenic variants in DNA damage repair genes were acquired through autosomal inheritance. Our study provides new insight into germline risk factors contributing to Ewing sarcoma pathogenesis.

## Introduction

Ewing sarcoma is the second most common bone and soft tissue cancer impacting children and adolescents worldwide^1^. It is an aggressive malignancy that is metastatic in 25% of cases at presentation and requires a very intensive treatment regimen including multiple chemotherapies, as well as surgery or radiation for local control. While overall survival for localized disease has improved to 75%, treatment confers significant morbidity, and cure rates for metastatic and relapsed disease remain poor^2^. Through a better understanding of the predisposing genetic factors contributing to Ewing sarcoma pathogenesis, the pediatric oncology community would be able to develop more informed and less toxic treatment regimens, as well as better screen children at risk for disease, opening the door to opportunities for earlier detection and even prevention.

Ewing sarcoma is driven by *EWSR1-ETS* gene fusions^1,3^, and is often characterized by a complex rearrangement pattern known as chromoplexy^4^. The genetic events preceding these simple and complex rearrangements remain largely unknown. Prior work has suggested a role for pathogenic germline variants in DNA damage repair (DDR) genes in Ewing sarcoma, but systematic case-control analyses to precisely define this role have not been undertaken^5,6^.

Much of what is known about germline predisposition to Ewing sarcoma has centered on common population variants identified as susceptibility loci from genome-wide association studies (GWAS)^1,7–9^, and a comprehensive evaluation of the relative contribution of rare coding pathogenic germline variants is largely incomplete.

Furthermore, a more complete understanding of the familial inheritance patterns of genetic risk factors in Ewing sarcoma is needed to guide cascade testing strategies with broad potential clinical impact. While guidelines for familial testing have been developed for various cancer predisposition syndromes^10,11^, patients with Ewing sarcoma and family members are not uniformly referred for genetic testing. Case reports of siblings with metachronous Ewing sarcoma diagnoses have suggested that germline variants shared within families may increase risk, but these have not yet been identified^12^. Family-based germline sequencing, such as the analysis of parent-proband trios, is thus a powerful tool for better understanding the inheritance of pathogenic germline variants in pediatric sarcoma generally, and Ewing sarcoma in particular^13^.

We hypothesized that through a systematic comparative analysis of germline predisposition across pediatric sarcoma subtypes, we would elucidate distinct patterns of rare coding pathogenic variants in Ewing sarcoma. We therefore undertook a three-stage study comprising 1) an ancestry-matched case-control analysis utilizing a discovery pan-sarcoma cohort, 2) validation with an ancestry-matched case-control analysis of an additional cohort of patients with Ewing sarcoma, and 3) evaluation of inheritance in germline variants for patients with Ewing sarcoma and their parents.

## Materials and Methods

### Ethics Approval and Consent to Participate

Written informed consent from patients and institutional review board approval, allowing comprehensive genetic analysis of germline samples, were obtained by the original studies that enrolled patients. The secondary genomic and deep-learning analyses performed for this study were approved under Dana-Farber Cancer Institute institutional review board protocols 21-143 and 20-691. This study conforms to the Declaration of Helsinki.

### Study Participants

A total of 1138 individuals with pediatric sarcoma diagnoses were included in the discovery cohort (Supplemental Methods). A combination of germline whole-genome sequencing (WGS) and whole-exome sequencing (WES) was aggregated for these individuals across four data sources; WGS was converted to WES equivalents using predefined target intervals to focus on coding variants only (Supplemental Methods). For validation, germline WGS for 425 individuals with Ewing sarcoma from the Gabriella Miller Kids First (GMKF) program was utilized. For 301 individuals with Ewing sarcoma from GMKF, germline WGS for parents was available (602 parents) and used for analysis of inheritance among trios (Tables S1 and S2; Supplemental Methods). Sequenced exomes for 24128 cancer-free individuals from six cohorts were extensively quality controlled, identically processed, and analyzed in the same way as cases for use as controls in this study (Supplemental Methods).

### Population Stratification

Principal component analysis (PCA) was undertaken utilizing germline genotypes for all discovery, validation, and control cohorts to enable ancestry inference. We used a trained random forest classifier to assign one of the five 1000 Genomes defined super populations (European, African, Admixed American, East Asian, and South Asian) to each sample in our case and control cohorts. Cases and controls were matched on genetic ancestry composition based on the first ten principal components from the preceding analysis (Supplemental Methods).

### Germline Variant Characterization

We called germline variants with a deep learning method, DeepVariant, which has shown superior sensitivity and specificity compared with a joint genotyping-based approach (version 0.8.0)^14,15^. High-quality coding variants were utilized for subsequent analyses (Supplemental Methods).

### Gene Sets

We evaluated the prevalence of pathogenic variants in a list of established germline cancer predisposition genes (n = 141; Table S3)^16–19^. A subset of these genes had an established role in DDR (n = 43). The low-penetrance founder *CHEK2* variant (p.Ile200Thr) was considered separately from other *CHEK2* pathogenic variants. We also evaluated predefined and non-mutually exclusive functional subsets of the DDR gene list for pathway-based enrichment analysis, curated through evaluation of known primary biological function in the Online Mendelian Inheritance in Man (OMIM)^20^ and Reactome^21^ databases: mismatch repair (n = 4), Fanconi Anemia (n = 16), double-strand break repair (n = 12), and nucleotide excision repair (n = 6).

### Germline Variant Pathogenicity Evaluation

Based on ClinVar database and Variant Effect Predictor (VEP) consequence annotations, all detected germline variants in cancer predisposition genes were classified into five categories: benign, likely benign, variants of unknown significance, likely pathogenic, and pathogenic using the American College of Medical Genetics (ACMG) guidelines^22^. Only putative loss-of-function, pathogenic, and likely pathogenic variants were included in this study (hereafter collectively referred to as pathogenic variants). Pathogenic variants were manually evaluated using the raw genomic data and the Integrative Genomics Viewer (IGV; Supplemental Methods; Tables S4, S5, and S6)^23,24^.

### Outcomes

The primary outcomes included gene-level enrichment analysis of germline pathogenic variants in individuals with Ewing sarcoma compared to other pediatric sarcomas, validation of enrichment findings in Ewing sarcoma, and analysis of mechanisms of inheritance among germline pathogenic variants in Ewing sarcoma. The secondary outcomes included exploratory gene and pathway level enrichment analysis of germline pathogenic variants in DDR genes in Ewing sarcoma.

### Statistical Analysis

Two-sided Fisher’s exact tests were used to calculate the odds ratios, 95% confidence intervals (CIs), and P values of germline pathogenic variant enrichment in affected versus unaffected cohorts for each of the examined cancer predisposition genes. P < 0.05 was the threshold for nominal enrichment signal. For the discovery cohort, the false discovery rate (FDR) was calculated using the Benjamini-Hochberg procedure; FDR < 0.05 was used as the threshold for enrichment meeting multiple hypothesis testing criteria for validation in the absence of a secondary cohort (applied to the discovery cohort; Supplemental Methods).

## Results

### Study overview and characteristics of discovery and validation cohorts

Our discovery cohort of 1138 primarily pediatric patients with sarcoma comprised osteosarcoma (436 cases), rhabdomyosarcoma (180 cases), Ewing sarcoma (222 cases), and other subtypes (300 cases; Figure 1A). The mean age of patients in the discovery cohort was 10.8 years (SD 5.5 years), and 52% of patients were male (Figure 1B). Our validation cohort comprised 433 patients with Ewing sarcoma; of these, 425 cases were available for an ancestry-matched case-control study, and 301 cases were available as parent-proband trios for evaluation for mechanisms of inheritance (Figure 1A). The mean age of patients in the Ewing sarcoma validation cohort was 13.3 years (SD 6.6 years), and 54% of patients were male (Figure 1C). The germline exome-wide mean target coverage for the discovery cohort samples was 53.9X (interquartile range [IQR] 37.6 – 66.3X), and for the Ewing sarcoma validation cohort samples was 27.3X (IQR 24.6 – 30.2X); exome-wide variant call rates were satisfactory for all samples (Figure S1). Differential coverage for evaluated genes was assessed, and overall comparable between cases and controls (Figure S2). All samples had satisfactory indel rates, variant transition-to-transversion rates, and genotype quality (Figure S3).

**Figure 1:**
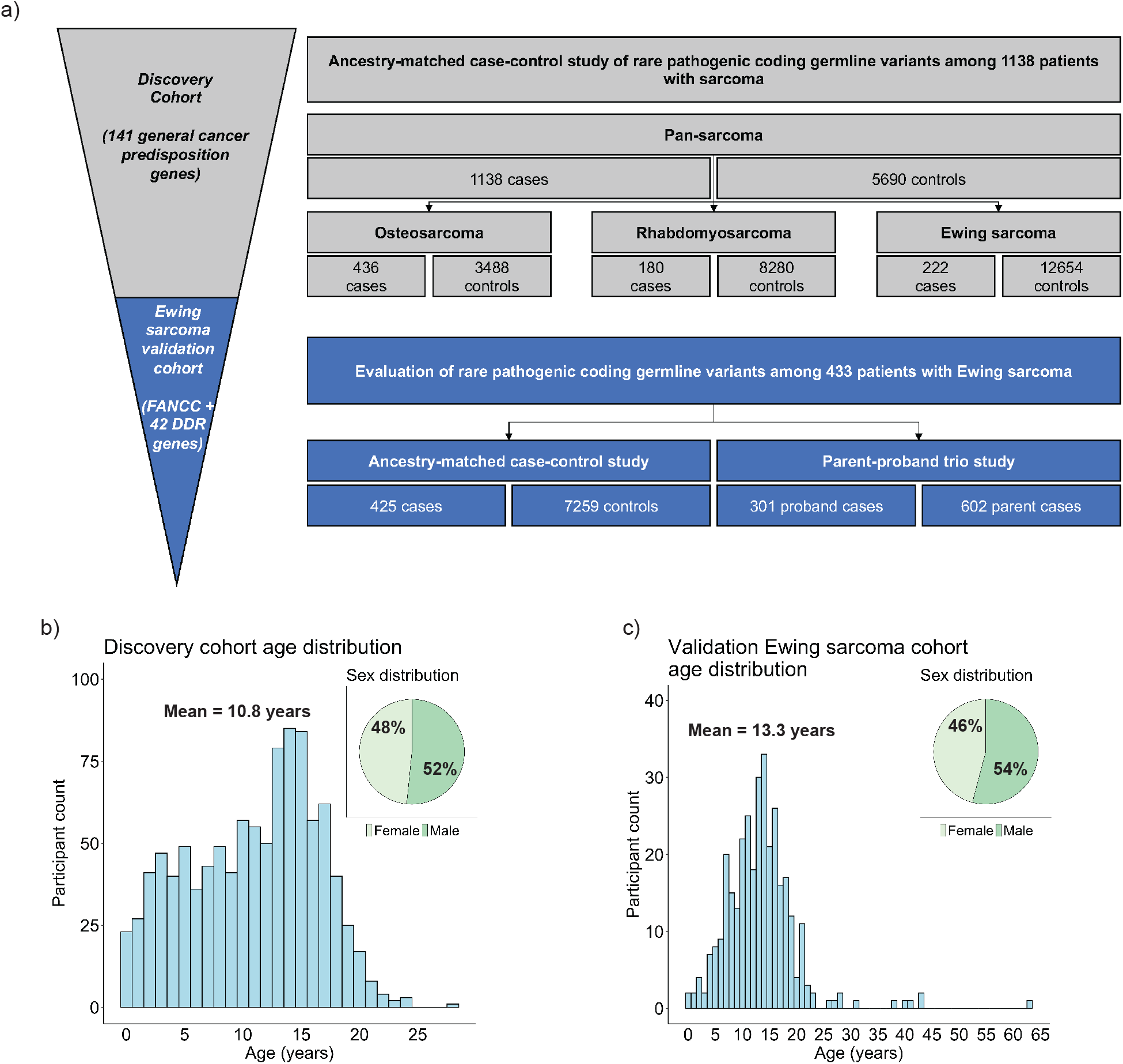
Study overview and characteristics of discovery and validation cohorts. A, Study schematic overview. The discovery cohort comprised 1138 cases, and the enrichment of pathogenic germline variants across 141 established cancer predisposition genes was evaluated. Ancestry-matched case-control analyses were carried out across the pan-sarcoma cohort (1138 cases), as well as major sarcoma histologic subtypes: osteosarcoma (436 cases), rhabdomyosarcoma (180 cases), Ewing sarcoma (222 cases). The validation cohort comprised 433 cases of Ewing sarcoma. The enrichment of pathogenic germline variants in 43 DNA damage repair genes was evaluated employing an ancestry-matched case-control analysis. Mechanisms of inheritance were evaluated for 301 cases available as parent-proband trios. B, Discovery cohort demographics: mean age 10.8 years, 52% male. C, Ewing sarcoma validation cohort demographics: mean age 13.3 years, 54% male.

### Pathogenic germline variants in cancer predisposition genes are enriched across pediatric sarcoma histologic subtypes relative to cancer-free controls

We assessed the frequency of pathogenic germline variants in 141 established cancer predisposition genes in our discovery cohort^16–19^. Our discovery cohort had broad representation from five major continental ancestries (Figure 2A; Figure S4); control cohorts for comparison were identically processed and ancestry-matched. The presence of pathogenic germline variants was not significantly associated with age (mean 10.9 vs. 10.5 years, p = 0.42 by two-sided t-test) or sex (p = 0.81 by Fisher’s exact test). Across the pan-sarcoma discovery cohort, nominal enrichment signal at p < 0.05 was observed for multiple genes previously implicated in sarcoma pathogenesis, including *TP53, NF1*, and *DICER1*^5,18,25^; nominal enrichment signal was also seen for *MUTYH, PALB2, NTHL1*, and *FANCC*, genes with less prior supporting evidence for their role in sarcoma germline predisposition. The enrichment in *TP53* was greatest, reaching significance at FDR < 0.05 across the pan-sarcoma discovery cohort (Figure 2B; Table S7).

**Figure 2:**
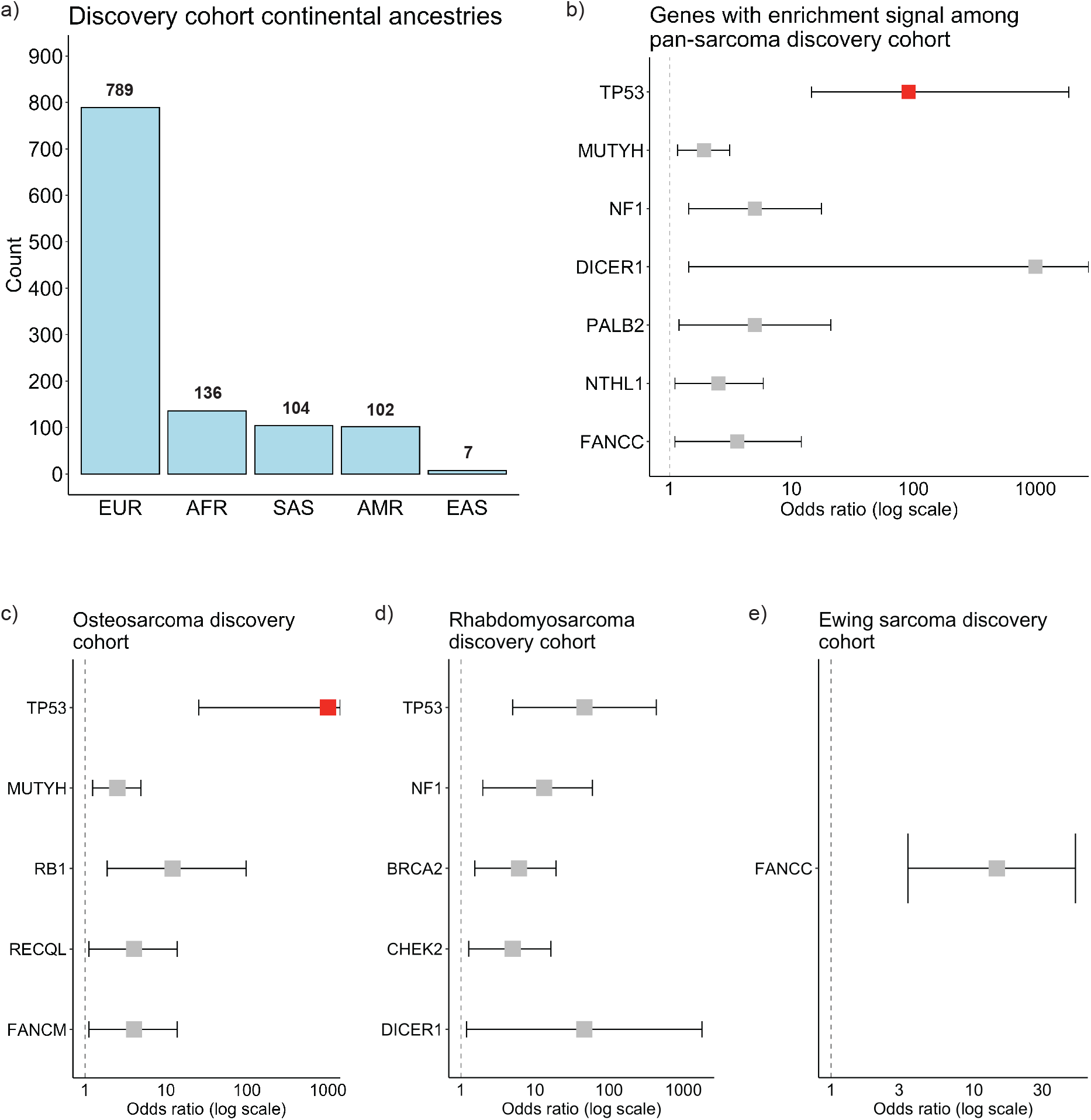
Ancestry composition of discovery cohort. Enrichment of pathogenic germline variants in discovery cohort, across and within major histologic subtypes. A, Ancestry composition of discovery cohort: 789 European cases (EUR), 136 African cases (AFR), 104 South Asian cases (SAS), 102 Admixed American cases (AMR), 7 East Asian cases (EAS). B – D, Odds ratios and 95% confidence intervals for enrichment of pathogenic germline variants among cancer predisposition genes. Red: Significant at FDR < 0.05 after Benjamini-Hochberg procedure. Gray: Significant at p < 0.05, but does not meet FDR < 0.05. Genes with p > 0.05 are not displayed. B, Pan-sarcoma cohort (1138 cases, 5690 controls). C, Osteosarcoma subset of cohort (436 cases, 3488 controls). D, Rhabdomyosarcoma subset of cohort (180 cases, 8280 controls). E, Ewing sarcoma subset of cohort (222 cases, 12654 controls).

We next carried out ancestry-matched enrichment analyses for each of the three major sarcoma histologic subtypes. For osteosarcoma, nominal enrichment signal was observed for 5 genes (Table S8). These included *TP53* and *RB1*, previously validated as important in germline predisposition to osteosarcoma (Figure 2C)^26^; once again, only *TP53* reached significance at FDR < 0.05. *RECQL* also had nominal enrichment signal in osteosarcoma cases relative to controls; however, due to relative underpowering, the previously implicated predisposition gene *RECQL4* did not, despite a higher absolute frequency in cases relative to controls (Figure S5). Nominal enrichment signal was also observed for *MUTYH* and *FANCM*, genes without substantial prior evidence supporting their role in germline predisposition to osteosarcoma^27^.

In rhabdomyosarcoma, nominal enrichment signal was observed for *TP53, NF1*, and *DICER1*, genes that have previously been implicated in germline predisposition to pediatric rhabdomyosarcoma^28^. We were able to redemonstrate a nominal enrichment signal for *BRCA2* and *CHEK2*, genes with a previous moderate level of evidence for a role in germline predisposition to rhabdomyosarcoma^25,29–31^. No genes reached significance at FDR < 0.05 (Figure 2D; Table S9).

In Ewing sarcoma, the only gene with nominal enrichment signal was *FANCC*, with heterozygous pathogenic variants seen in 3 out of 222 cases (1.3%, OR 14.4, 95% CI 3.5 – 51.2, p = 0.002, FDR = 0.28; Figure 2E; Figure S5; Table S10). Prior work had shown a general association between germline variants in Fanconi Anemia pathway genes and translocation-driven sarcomas, but the role of *FANCC* in germline predisposition to Ewing sarcoma had not previously been reported to our knowledge^5^. In contrast to osteosarcoma and rhabdomyosarcoma, no pathogenic germline *TP53* variants were observed in Ewing sarcoma.

Thus, our enrichment analysis of pathogenic variants in our discovery cohort demonstrated a unique pattern of predisposing variants across pediatric sarcoma subtypes, with a strong enrichment signal for *TP53* in all sarcoma subtypes except Ewing sarcoma. Having demonstrated a distinct enrichment pattern among Ewing sarcoma cases relative to osteosarcoma and rhabdomyosarcoma, we proceeded to evaluate the enrichment signal in *FANCC* in our larger validation cohort of Ewing sarcoma cases.

### FANCC and other DNA damage repair genes harbor pathogenic germline variants in Ewing sarcoma

Our validation cohort of Ewing sarcoma patients was also ancestry-matched to cancer-free controls to enable targeted evaluation of *FANCC* enrichment (Figure S6). Heterozygous pathogenic germline *FANCC* variants were again enriched, seen in 3 of 425 cases (0.7%, OR 5.1, 95% CI 1.2 – 18.5, p = 0.03, single hypothesis; Figure 3A). The pooled odds ratio for *FANCC* enrichment between discovery and validation cohorts remained significant (6 of 647 cases, 0.9%, OR 7.7, 95% CI 3.1 – 19.3, p < .0001).

**Figure 3:**
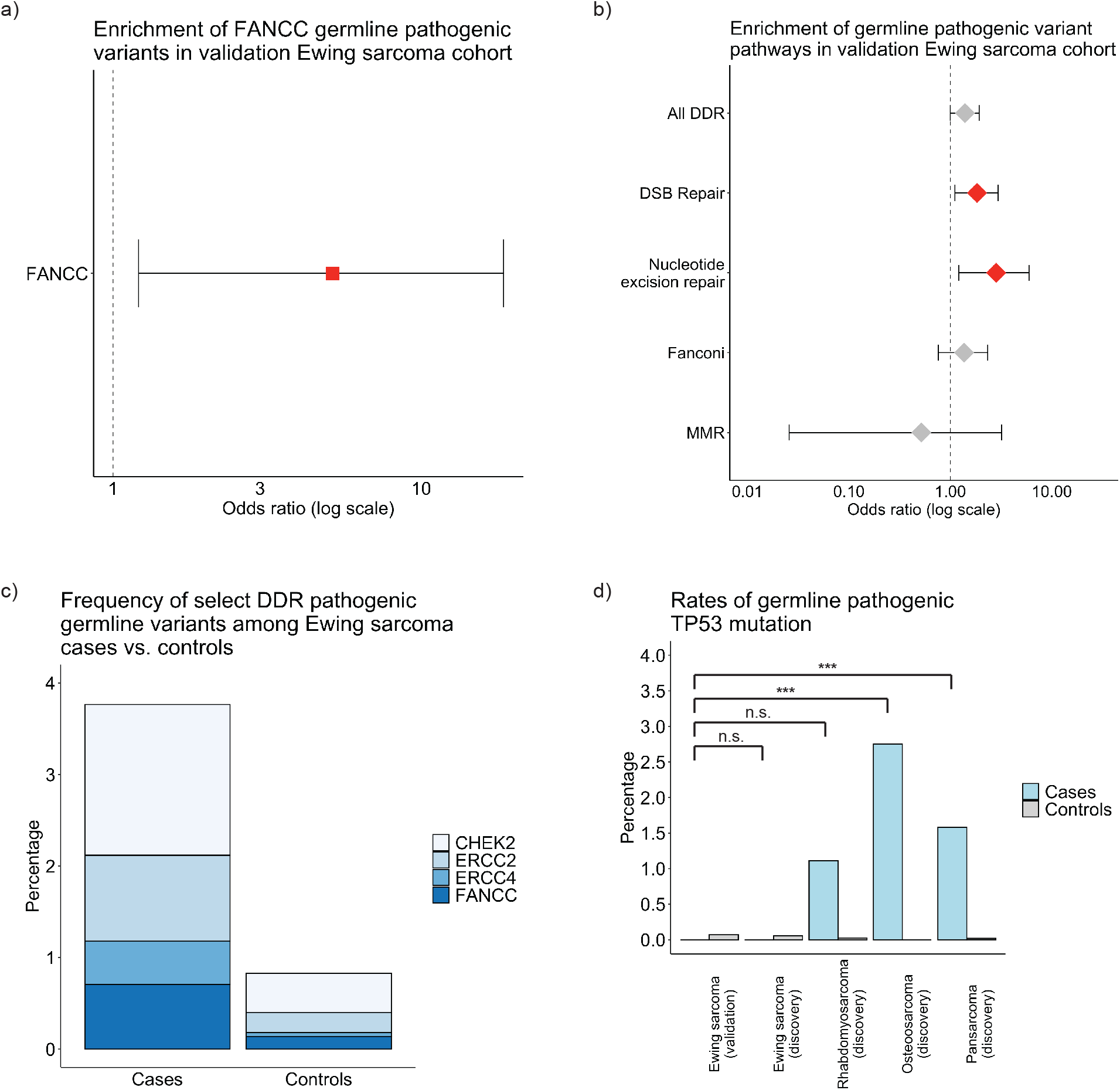
Enrichment of pathogenic germline variants in *FANCC* and other DNA damage repair genes in Ewing sarcoma validation cohort. A, Enrichment of pathogenic germline variants in *FANCC* in the Ewing Sarcoma validation cohort vs. controls (OR 5.1, 95% CI 1.2 – 18.5). B, Enrichment of pathogenic germline variants in DNA damage response genes and specific pathways (Red: Significant at p < 0.05; Gray: Not significant): 43 DNA damage response genes (All DDR; OR 1.4, 95% CI 1.0 – 1.9, p = 0.05), 12 genes involved in DNA double-strand break repair (DSB repair; OR 1.8, 95% CI 1.1 – 3.0, p = 0.02), 6 genes involved in nucleotide excision repair (OR 2.8, 95% CI 1.2 – 5.9, p = 0.01), 16 genes involved in Fanconi anemia pathway (OR 1.4, 95% CI 0.8 – 2.3, p = 0.28), 4 genes involved in mismatch repair (MMR; OR 0.5, 95% CI 0 – 3.2, p = 1.0). C, Collective frequency of the 4 genes with enrichment signal at p < 0.05 (*CHEK2, ERCC2, ERCC4*, and *FANCC*) in Ewing sarcoma cases vs. controls. D, Rates of pathogenic variants in *TP53* in Ewing sarcoma validation cohort in comparison to Ewing sarcoma subset of discovery cohort, rhabdomyosarcoma subset of discovery cohort, osteosarcoma subset of discovery cohort, and pan-sarcoma discovery cohort (Fisher’s exact tests, n.s. denotes no significant difference, *** denotes significant difference at p < 0.05).

Prior mechanistic work has demonstrated that *FANCC* knockout contributes to rearrangement signatures consistent with homologous recombination deficiency^32^. We thus asked whether the recurrent enrichment of *FANCC* represented a broader importance of DDR genes in germline predisposition to Ewing sarcoma, as has been previously suggested^6^. We performed an exploratory analysis in the Ewing sarcoma validation cohort on the subset of 43 cancer predisposition genes with specific DDR pathway roles. In aggregate, pathogenic variants in DNA double-strand break repair (DSB) genes (OR 1.8, 95% CI 1.1 – 3.0, p = 0.02) and nucleotide excision repair genes (OR 2.8, 95% CI 1.2 – 5.9, p = 0.01; Figure 3B, Table S11) were enriched. While the discovery Ewing sarcoma and rhabdomyosarcoma cohorts were underpowered for a comparative pathway analysis (Tables S12 and S13), the discovery osteosarcoma cohort was of comparable size. In aggregate, DDR genes were enriched in discovery osteosarcoma cases (OR 1.9, 95% CI 1.4 – 2.6, p = 0.0001, Figure S7); notably and in contrast to Ewing sarcoma, this was driven largely by germline *TP53* mutations, as the DSB and nucleotide excision repair pathways were not significantly enriched (Table S14). In the validation cohort of patients with Ewing sarcoma, the presence of pathogenic germline variants in DDR genes was not significantly associated with age (mean 13.3 vs. 13.5 years, p = 0.88 by two-sided t-test) or sex (p = 0.10 by Fisher’s exact test).

In evaluating the specific genes contributing to the collective enrichment of the DSB and nucleotide excision repair pathways, we found that *CHEK2, ERCC2*, and *ERCC4* had nominal enrichment signal at p < 0.05 in our validation Ewing sarcoma cohort, although none of these individual genes reached significance at FDR < 0.05 when correcting for 43 DDR genes (Table S15). In combination with *FANCC*, these four genes harbored germline pathogenic variants in 16 of 425 Ewing sarcoma cases (3.8%), compared with 60 of 7259 controls (0.8%; Figure 3C).

Similar to our discovery Ewing sarcoma cohort, we once again identified no pathogenic germline *TP53* variants in our validation Ewing sarcoma cohort. The rate of pathogenic germline *TP53* variants in patients with Ewing sarcoma (0%) was significantly lower than that seen for all sarcomas in aggregate (1.6%, p = 0.006 by Fisher’s exact test), and osteosarcoma in particular (2.8%, p = 0.0005 by Fisher’s exact test; Figure 3D). This, in combination with the recurrent enrichment of pathogenic germline mutations in *FANCC*, as well as the collective enrichment of DSB and nucleotide excision repair pathways, demonstrated a distinct pattern of germline mutations in Ewing sarcoma relative to other pediatric sarcoma subtypes.

### Pathogenic germline variants in DNA damage repair genes are inherited in high-risk families

Having identified pathogenic germline mutations in *FANCC* and other DDR genes in Ewing sarcoma, we next sought to assess inheritance of these variants. Thus, we evaluated the 301 patients with Ewing sarcoma from our validation cohort that were part of parent-proband sequencing trios. Among these 301 patients, 32 harbored pathogenic germline variants in DDR genes (10.6%; Figure 4A). In 32 of 32 probands in which a pathogenic germline DDR variant was identified in a proband, the same germline DDR variant was identified in one of the parents (100%). In contrast, for probands in whom a pathogenic germline DDR variant was not identified, only 19 of 269 had at least one parent with a germline DDR variant (7.1%; Figure 4B). Collectively, pathogenic variants in DSB repair genes were more common among parents of probands than ancestry-matched cancer-free controls (OR 1.8, 95% CI 1.2 – 2.7, p = 0.006). While pathogenic germline variants in genes such as *BRCA2* and *CHEK2* were also observed in some parents and not inherited by probands, these were at a rate that was comparable to the population frequency (Figure S8).

**Figure 4:**
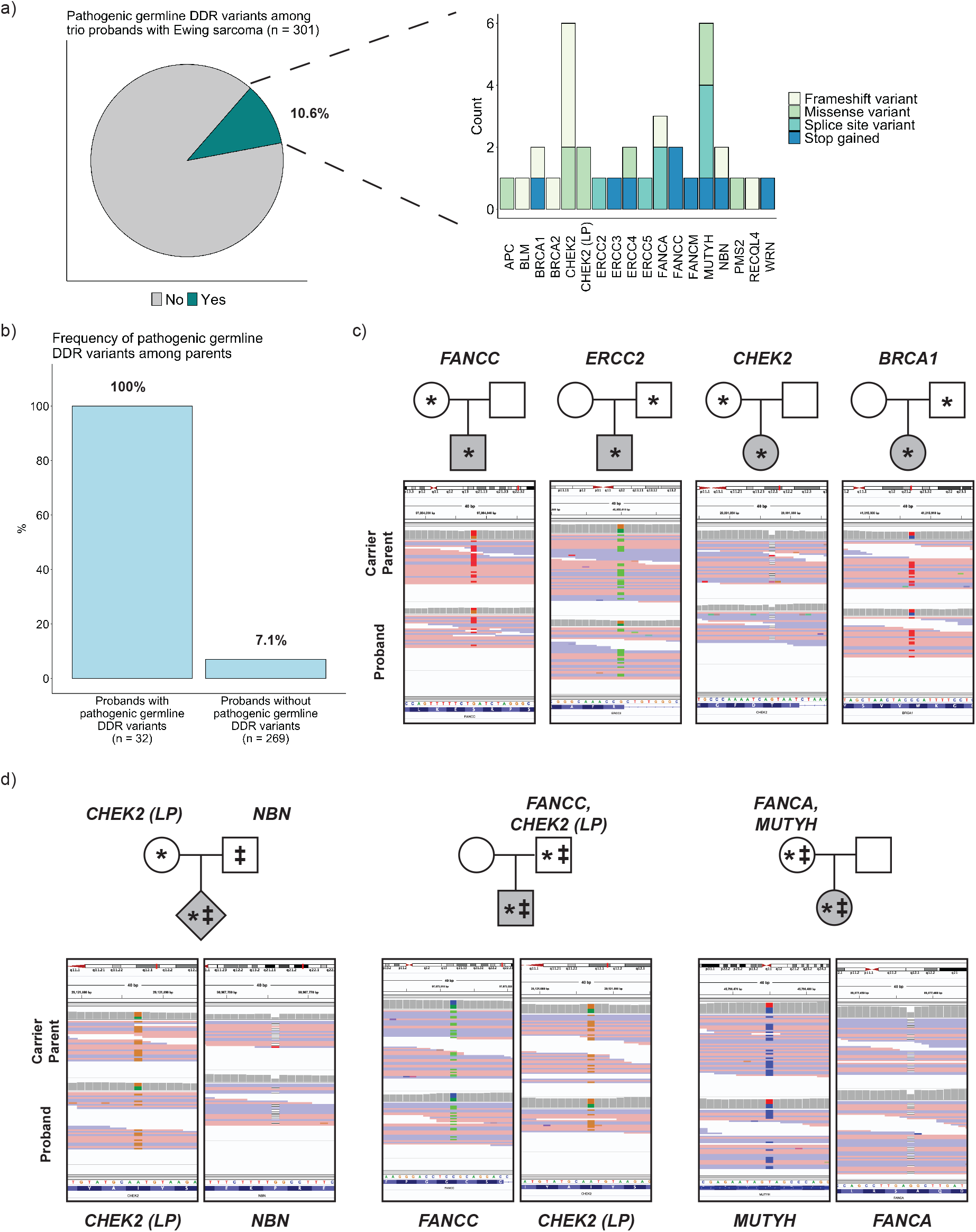
Pathogenic germline variants in DNA damage repair genes are inherited in high-risk families. A, Pathogenic germline variants in DDR genes among trio probands with Ewing sarcoma. 35 pathogenic variants impacting 32 of 301 patients with Ewing sarcoma were identified. B, 32 of 32 probands (100%) with pathogenic germline variants in DDR genes had identical variants identified in parents. 19 of 269 probands without a pathogenic germline variant in a DDR gene had at least one parent with a germline DDR variant that was not inherited by the proband (7.1%). C – D, Pedigrees and IGV screenshots of pathogenic variants in DDR genes. Pedigree legend: circle = female sex, square = male sex, diamond = unknown sex, gray shading = proband with Ewing sarcoma, * denotes variant 1 identified in parent-proband trio, ‡ denotes variant 2 identified in parent-proband trio. Top screenshot: Carrier Parent, Bottom screenshot: Proband.

Identical pathogenic germline DDR variants in probands and parents impacted *FANCC, ERCC2, CHEK2*, and *BRCA1* among other genes (Figure 4C). In three instances, heterozygous germline pathogenic variants affecting multiple DDR genes were seen in probands, with each variant also identified in a parent (Figure 4D).

We sought to understand whether as yet unidentified *de novo* pathogenic variants in other coding genes may coordinate with or complement the inherited DDR variants to explain a significant proportion of the unexplained germline risk for developing pediatric Ewing sarcoma. Based on prior frameworks^33–35^, we reasoned that finding pathogenic *de novo* variants recurrently impacting the same gene in a cohort of 301 proband-parent trios would be highly unlikely by chance, implicating potential additional candidate risk genes. However, in our cohort, we identified recurrent pathogenic *de novo* germline variants in only one gene, *TTN*, which occurred in two separate Ewing sarcoma cases. As the frequency of pathogenic germline variants in *TTN* between cases and cancer-free controls was not significantly different and there is no established biological role for *TTN* in Ewing sarcoma oncogenesis, there was insufficient evidence to support its role in germline predisposition to Ewing sarcoma (Figure S9).

Taken together, pathogenic germline variants in DSB genes were present more frequently in families of patients with Ewing sarcoma relative to cancer-free controls. Autosomal inheritance, as opposed to *de novo* development, was the mechanism of inheritance of these moderate penetrance risk variants.

## Discussion

This study represents the largest and most systematic analysis of germline predisposition to Ewing sarcoma relative to other pediatric sarcoma subtypes to date. Having assembled germline sequencing data from 1138 patients with pediatric sarcoma diagnoses, we undertook rigorous ancestry-matched case-control analyses and illustrated distinct patterns of enrichment amongst pathogenic variants in Ewing sarcoma relative to osteosarcoma and rhabdomyosarcoma. Supporting the validity of our approach, we were able to recover enrichment signal in many cancer predisposition genes known to be associated with pediatric sarcoma risk, such as *TP53, RB1*, and *DICER1*. We additionally demonstrated enrichment signal in several cancer predisposition genes with less well-characterized links to pediatric sarcoma, most notably *FANCC* in Ewing sarcoma. We then validated the enrichment of pathogenic germline variants in *FANCC* among patients with Ewing sarcoma using an independent cohort of 425 unique patients. This recurrent enrichment of heterozygous pathogenic germline variants in *FANCC* provides evidence for its role in increasing risk for some cases of Ewing sarcoma, and raises the possibility that monoallelic germline variants in Fanconi anemia genes may confer increased risk in other translocation-associated cancers.

We demonstrated that the enrichment in *FANCC* pathogenic germline variants represented a broader importance of DNA damage repair (DDR) genes, and both the DNA double-strand break (DSB) and nucleotide excision repair pathways in particular, for Ewing sarcoma germline predisposition. While prior studies have identified occasional instances of pathogenic germline *TP53* variants amongst cases of Ewing sarcoma, through comparative analyses, we found that the frequency of pathogenic germline *TP53* variants among cases of Ewing sarcoma was significantly lower in relation to other pediatric sarcoma subtypes^6,29^. This finding is supported by the clinical observation that Ewing sarcoma is not frequently seen in families with Li-Fraumeni syndrome^36^.

Using parent-proband trios, we showed that pathogenic germline variants in DNA damage repair genes found in patients with Ewing sarcoma are also present in their parents, and therefore passed on through autosomal inheritance. We found pathogenic germline variants in DSB genes enriched amongst parents of patients with Ewing sarcoma. As moderate penetrance risk variants that are also present in parents, we reasoned that pathogenic germline variants in DDR genes play a substantial role in increasing risk for developing Ewing sarcoma, but are likely not sufficient to cause the disease in isolation. However, we did not identify *de novo* variants recurrently impacting other genes to support a role for their interaction with pathogenic germline variants in DDR genes to promote germline predisposition to Ewing sarcoma.

Our study had some notable limitations. While pathogenic germline variants in *FANCC* occurred at a rate greater than expected by chance in cases of Ewing sarcoma, the overall frequency of these variants was low (1.3% in the discovery cohort, 0.7% in the validation cohort), supporting the role of *FANCC* as a moderate penetrance cancer predisposition gene as opposed to the sole driver of disease pathogenesis. Additionally, our focus on germline variants in select DDR genes within Ewing sarcoma likely underestimated the total contribution of rare coding pathogenic germline variants to Ewing sarcoma pathogenesis. Finally, similar to much preceding work in germline predisposition in pediatric cancers, our methods were limited to identifying known pathogenic germline SNVs/ indels conferring increased risk in pediatric sarcoma. As progress is made in germline structural variant discovery^37^ and placing rare pathogenic variants in the context of complex germline interactions^38–41^, future studies and new statistical frameworks will be needed to more completely define the role of germline predisposition in Ewing sarcoma pathogenesis.

Taken together, our analysis supports a unique contribution of germline variants in *FANCC* and other DDR genes to Ewing sarcoma pathogenesis. Our study provides a foundation to inform approaches to genetic testing for patients with Ewing sarcoma, as well as cascade testing for family members.

## Data Availability

All data produced in the present work are contained in the manuscript. Raw sequencing data is available through public data repositories as detailed in the section pertaining to source data.

## Acknowledgments

This work was supported by a Research Training Grant in Pediatric Oncology NIH 5T32CA136432-12 (R.G.), American Society of Clinical Oncology Conquer Cancer – Sarcoma Foundation of America Young Investigator Award (R.G.), Jay Vernon Jackson Memorial Research Award from Sarcoma Foundation of America (R.G.), Edward B. Clark, MD Endowed Chair in Pediatric Research (J.D.S.), Helen Clise Presidential Endowed Chair in Li-Fraumeni Syndrome Research (J.D.S.), Conquer Cancer Foundation Career Development Award 13167 from the American Society of Clinical Oncology (S.H.A.), Young Investigator Award 18YOUN02 from the Prostate Cancer Foundation (S.H.A.), Physician Research Award (PC200150/W81XWH-21-1-0084) of the Department of Defense (S.H.A), NIH R37 CA222574 (E.M.V.A.), R01 CA227388 (E.M.V.A.), U01 CA233100 (E.M.V.A.), Innovation in Cancer Informatics Award (E.M.V.A.), COG Biospecimen Bank Grant U24CA196173, NCTN Operations Center Grant U10CA180886, NCTN Statistics & Data Center Grant U10CA180899, and Gabriella Miller Kids First 1×01 HL 140547-01.

## Data and code availability

Sequencing data analyzed in this study were obtained from multiple publicly accessible data repositories. Sequenced samples from the discovery cohort came from four data sources: St. Jude Cloud (Pediatric Cancer Genome Project, St. Jude Lifetime, Genomes for Kids, and Childhood Cancer Survivor Study; n = 1033)^42–46^, dbGaP study “Genomic Sequencing of Ewing Sarcoma” (phs000804.v1.p1; n = 26)^47,48^, dbGaP study “Osteosarcoma Genomics” (phs000699.v1.p1, n = 58)^49^, and ICGC study “Bone Cancer – UK” (BOCA-UK, n = 63)^50^. Sequenced samples from the Ewing sarcoma validation and trio cohorts were from the Gabriella Miller Kids First “Ewing Sarcoma – Genetic Risk” study (phs001228.v1.p1)^51^. The control cohorts came from the following sources: Autism Sequencing Consortium (dbGaP phs000298), Framingham Cohort (dbGaP phs000007), Multi-Ethnic Study of Atherosclerosis (dbGaP phs000209), Lung Cohort (dbGaP phs000291), in-house collection of exomes from National Heart, Lung, and Blood Institute “Grand Opportunity” Exome Sequencing Project (NHLBI GO-ESP), and the 1000 Genomes Project^52^.

## Supplemental data

Supplemental data includes supplemental methods, 9 supplemental figures, and 15 supplemental tables.

## Author contributions

R.G., E.M.V.A., and S.H.A. conceived the project design, carried out the analysis, and wrote the manuscript. B.D.C. and K.A.J. contributed to refinement of the project design, data interpretation, validation of findings, and manuscript review. S.Y.C., S.A., and J.K.J. provided support with data analysis, data interpretation, and manuscript review. J.D.C. contributed to validation of findings, data interpretation, and manuscript review. E.Y., S.O., L.H., G.M., and T.F. carried out foundational work that enabled validation of findings. J.K. and A.G. contributed to data interpretation and manuscript review.

## Disclaimer

The content is solely the responsibility of the authors and does not necessarily represent the official views of the National Institutes of Health.

## References

1. Grünewald, T.G.P., Cidre-Aranaz, F., Surdez, D., Tomazou, E.M., De Álava, E., Kovar, H., Sorensen, P.H., Delattre, O., and Dirksen, U. (2018). Ewing sarcoma. Nat. Rev. Dis. Prim. 4,.

2. Balamuth, N.J., and Womer, R.B. (2010). Ewing’s sarcoma. Lancet Oncol. 11, 184–192.

3. Brohl, A.S., Solomon, D.A., Chang, W., Wang, J., Song, Y., Sindiri, S., Patidar, R., Hurd, L., Chen, L., Shern, J.F., et al. (2014). The Genomic Landscape of the Ewing Sarcoma Family of Tumors Reveals Recurrent STAG2 Mutation. 10,.

4. Anderson, N.D., De Borja, R., Young, M.D., Fuligni, F., Rosic, A., Roberts, N.D., Hajjar, S., Layeghifard, M., Novokmet, A., Kowalski, P.E., et al. (2018). Rearrangement bursts generate canonical gene fusions in bone and soft tissue tumors. Science (80-.). 361,.

5. Ballinger, M.L., Goode, D.L., Ray-Coquard, I., James, P.A., Mitchell, G., Niedermayr, E., Puri, A., Schiffman, J.D., Dite, G.S., Cipponi, A., et al. (2016). Monogenic and polygenic determinants of sarcoma risk: an international genetic study. Lancet Oncol. 17, 1261–1271.

6. Brohl, A.S., Patidar, R., Turner, C.E., Wen, X., Song, Y.K., Wei, J.S., Calzone, K.A., and Khan, J. (2017). Frequent inactivating germline mutations in DNA repair genes in patients with Ewing sarcoma. Genet. Med. 19, 955–958.

7. Grünewald, T.G.P., Bernard, V., Gilardi-Hebenstreit, P., Raynal, V., Surdez, D., Aynaud, M.M., Mirabeau, O., Cidre-Aranaz, F., Tirode, F., Zaidi, S., et al. (2015). Chimeric EWSR1-FLI1 regulates the Ewing sarcoma susceptibility gene EGR2 via a GGAA microsatellite. Nat. Genet. 47, 1073–1078.

8. Postel-Vinay, S., Véron, A.S., Tirode, F., Pierron, G., Reynaud, S., Kovar, H., Oberlin, O., Lapouble, E., Ballet, S., Lucchesi, C., et al. (2012). Common variants near TARDBP and EGR2 are associated with susceptibility to Ewing sarcoma. Nat. Genet. 44, 323–327.

9. Machiela, M.J., Grünewald, T.G.P., Surdez, D., Reynaud, S., Mirabeau, O., Karlins, E., Rubio, R.A., Zaidi, S., Grossetete-Lalami, S., Ballet, S., et al. (2018). Genome-wide association study identifies multiple new loci associated with Ewing sarcoma susceptibility. Nat. Commun. 9, 1–8.

10. Kratz, C.P., Achatz, M.I., Brugieres, L., Frebourg, T., Garber, J.E., Greer, M.L.C., Hansford, J.R., Janeway, K.A., Kohlmann, W.K., McGee, R., et al. (2017). Cancer screening recommendations for individuals with Li-Fraumeni syndrome. Clin. Cancer Res. 23, e38–e45.

11. Schultz, K.A.P., Williams, G.M., Kamihara, J., Stewart, D.R., Harris, A.K., Bauer, A.J., Turner, J., Shah, R., Schneider, K., Schneider, K.W., et al. (2018). Dicer1 and associated conditions: Identification of at-risk individuals and recommended surveillance strategies. Clin. Cancer Res. 24, 2251–2261.

12. Joyce, M.J., Harmon, D.C., Mankin, H.J., Suit, H.D., Schiller, A.L., and Truman, J.T. (1984). Ewing’s sarcoma in female siblings: A clinical report and review of the literature. Cancer 53, 1959–1962.

13. Kuhlen, M., Taeubner, J., Brozou, T., Wieczorek, D., Siebert, R., and Borkhardt, A. (2019). Family-based germline sequencing in children with cancer. Oncogene 38, 1367–1380.

14. Poplin, R., Chang, P.C., Alexander, D., Schwartz, S., Colthurst, T., Ku, A., Newburger, D., Dijamco, J., Nguyen, N., Afshar, P.T., et al. (2018). A universal snp and small-indel variant caller using deep neural networks. Nat. Biotechnol. 36, 983.

15. AlDubayan, S.H., Conway, J.R., Camp, S.Y., Witkowski, L., Kofman, E., Reardon, B., Han, S., Moore, N., Elmarakeby, H., Salari, K., et al. (2020). Detection of pathogenic variants with germline genetic testing using deep learning vs standard methods in patients with prostate cancer and melanoma. JAMA - J. Am. Med. Assoc. 324, 1957–1969.

16. Rahman, N. (2014). Realizing the promise of cancer predisposition genes. Nature 505, 302–308.

17. Huang, K. lin, Mashl, R.J., Wu, Y., Ritter, D.I., Wang, J., Oh, C., Paczkowska, M., Reynolds, S., Wyczalkowski, M.A., Oak, N., et al. (2018). Pathogenic Germline Variants in 10,389 Adult Cancers. Cell 173, 355–370.e14.

18. Mirabello, L., Zhu, B., Koster, R., Karlins, E., Dean, M., Yeager, M., Gianferante, M., Spector, L.G., Morton, L.M., Karyadi, D., et al. (2020). Frequency of Pathogenic Germline Variants in Cancer-Susceptibility Genes in Patients with Osteosarcoma. JAMA Oncol. 6, 724–734.

19. Tate, J.G., Bamford, S., Jubb, H.C., Sondka, Z., Beare, D.M., Bindal, N., Boutselakis, H., Cole, C.G., Creatore, C., Dawson, E., et al. (2019). COSMIC: The Catalogue Of Somatic Mutations In Cancer. Nucleic Acids Res. 47, D941–D947.

20. Amberger, J.S., Bocchini, C.A., Schiettecatte, F., Scott, A.F., and Hamosh, A. (2015). OMIM.org: Online Mendelian Inheritance in Man (OMIM®), an Online catalog of human genes and genetic disorders. Nucleic Acids Res. 43, D789–D798.

21. Jassal, B., Matthews, L., Viteri, G., Gong, C., Lorente, P., Fabregat, A., Sidiropoulos, K., Cook, J., Gillespie, M., Haw, R., et al. (2020). The reactome pathway knowledgebase. Nucleic Acids Res. 48, D498–D503.

22. Richards, S., Aziz, N., Bale, S., Bick, D., Das, S., Gastier-Foster, J., Grody, W.W., Hegde, M., Lyon, E., Spector, E., et al. (2015). Standards and guidelines for the interpretation of sequence variants: A joint consensus recommendation of the American College of Medical Genetics and Genomics and the Association for Molecular Pathology. Genet. Med. 17, 405–424.

23. Robinson, J.T., Thorvaldsdóttir, H., Winckler, W., Guttman, M., Lander, E.S., Getz, G., and Mesirov, J.P. (2011). Integrative genomics viewer. Nat. Biotechnol. 29, 24–26.

24. Robinson, J.T., Thorvaldsdóttir, H., Wenger, A.M., Zehir, A., and Mesirov, J.P. (2017). Variant review with the integrative genomics viewer. Cancer Res. 77, e31–e34.

25. Li, H., Sisoudiya, S.D., Martin-Giacalone, B.A., Khayat, M.M., Dugan-Perez, S., Marquez-Do, D.A., Scheurer, M.E., Muzny, D., Boerwinkle, E., Gibbs, R.A., et al. (2020). Germline Cancer Predisposition Variants in Pediatric Rhabdomyosarcoma: A Report From the Children’s Oncology Group. JNCI J. Natl. Cancer Inst. 00, 1–9.

26. Gianferante, D.M., Mirabello, L., and Savage, S.A. (2017). Germline and somatic genetics of osteosarcoma - Connecting aetiology, biology and therapy. Nat. Rev. Endocrinol. 13, 480–491.

27. Mirabello, L., Yu, K., Berndt, S.I., Burdett, L., Wang, Z., Chowdhury, S., Teshome, K., Uzoka, A., Hutchinson, A., Grotmol, T., et al. (2011). A comprehensive candidate gene approach identifies genetic variation associated with osteosarcoma. BMC Cancer 11,.

28. Martin-Giacalone, B.A., Weinstein, P.A., Plon, S.E., and Lupo, P.J. (2021). Pediatric Rhabdomyosarcoma: Epidemiology and Genetic Susceptibility. J. Clin. Med. 10, 2028.

29. Zhang, J., Walsh, M.F., Wu, G., Edmonson, M.N., Gruber, T.A., Easton, J., Hedges, D., Ma, X., Zhou, X., Yergeau, D.A., et al. (2015). Germline mutations in predisposition genes in pediatric cancer. N. Engl. J. Med. 373, 2336–2346.

30. Kim, J., Light, N., Subasri, V., Young, E.L., Wegman-Ostrosky, T., Barkauskas, D.A., Hall, D., Lupo, P.J., Patidar, R., Maese, L.D., et al. (2021). Pathogenic Germline Variants in Cancer Susceptibility Genes in Children and Young Adults With Rhabdomyosarcoma. JCO Precis. Oncol. 75–87.

31. Naslund-Koch, C., Nordestgaard, B.G., and Bojesen, S.E. (2016). Increased risk for other cancers in addition to breast cancer for CHEK2*1100delC heterozygotes estimated from the copenhagen general population study. J. Clin. Oncol. 34, 1208–1216.

32. Zou, X., Owusu, M., Harris, R., Jackson, S.P., Loizou, J.I., and Nik-Zainal, S. (2018). Validating the concept of mutational signatures with isogenic cell models. Nat. Commun. 9, 1–16.

33. Ba, W., Yan, Y., Reijnders, M.R.F., Schuurs-Hoeijmakers, J.H.M., Feenstra, I., Bongers, E.M.H.F., Bosch, D.G.M., De Leeuw, N., Pfundt, R., Gilissen, C., et al. (2016). TRIO loss of function is associated with mild intellectual disability and affects dendritic branching and synapse function. Hum. Mol. Genet. 25, 892–902.

34. Jin, Z.B., Wu, J., Huang, X.F., Feng, C.Y., Cai, X.B., Mao, J.Y., Xiang, L., Wu, K.C., Xiao, X., Kloss, B.A., et al. (2017). Trio-based exome sequencing arrests de novo mutations in early-onset high myopia. Proc. Natl. Acad. Sci. U. S. A. 114, 4219–4224.

35. Samocha, K.E., Robinson, E.B., Sanders, S.J., Stevens, C., Sabo, A., McGrath, L.M., Kosmicki, J.A., Rehnström, K., Mallick, S., Kirby, A., et al. (2014). A framework for the interpretation of de novo mutation in human disease. Nat. Genet. 46, 944–950.

36. Kratz, C.P., Freycon, C., Maxwell, K.N., Nichols, K.E., Schiffman, J.D., Evans, D.G., Achatz, M.I., Savage, S.A., Weitzel, J.N., Garber, J.E., et al. (2021). Analysis of the Li-Fraumeni Spectrum Based on an International Germline TP53 Variant Data Set: An International Agency for Research on Cancer TP53 Database Analysis. JAMA Oncol.

37. Collins, R.L., Brand, H., Karczewski, K.J., Zhao, X., Alföldi, J., Francioli, L.C., Khera, A. V., Lowther, C., Gauthier, L.D., Wang, H., et al. (2020). A structural variation reference for medical and population genetics. Nature 581, 444–451.

38. Fahed, A.C., Wang, M., Homburger, J.R., Patel, A.P., Bick, A.G., Neben, C.L., Lai, C., Brockman, D., Philippakis, A., Ellinor, P.T., et al. (2020). Polygenic background modifies penetrance of monogenic variants for tier 1 genomic conditions. Nat. Commun. 11, 1–9.

39. Wand, H., Lambert, S.A., Tamburro, C., Iacocca, M.A., O’Sullivan, J.W., Sillari, C., Kullo, I.J., Rowley, R., Dron, J.S., Brockman, D., et al. (2021). Improving reporting standards for polygenic scores in risk prediction studies. Nature 591, 211–219.

40. Sud, A., Turnbull, C., and Houlston, R. (2021). Will polygenic risk scores for cancer ever be clinically useful? Npj Precis. Oncol. 5, 1–5.

41. Seplyarskiy, V.B., Soldatov, R.A., Koch, E., McGinty, R.J., Goldmann, J.M., Hernandez, R.D., Barnes, K., Correa, A., Burchard, E.G., Ellinor, P.T., et al. (2021). Population sequencing data reveal a compendium of mutational processes in the human germ line. Science (80-.). eaba7408.

42. McLeod, C., Gout, A.M., Zhou, X., Thrasher, A., Rahbarinia, D., Brady, S.W., Macias, M., Birch, K., Finkelstein, D., Sunny, J., et al. (2021). St. Jude cloud: A pediatric cancer genomic data-sharing ecosystem. Cancer Discov. 11, 1082–1099.

43. Chen, X., Stewart, E., Shelat, A.A., Qu, C., Bahrami, A., Hatley, M., Wu, G., Bradley, C., McEvoy, J., Pappo, A., et al. (2013). Targeting Oxidative Stress in Embryonal Rhabdomyosarcoma. Cancer Cell 24, 710–724.

44. Downing, J.R., Wilson, R.K., Zhang, J., Mardis, E.R., Pui, C.H., Ding, L., Ley, T.J., and Evans, W.E. (2012). The pediatric cancer genome project. Nat. Genet. 44, 619–622.

45. Wang, Z., Wilson, C.L., Easton, J., Thrasher, A., Mulder, H., Liu, Q., Hedges, D.J., Wang, S., Rusch, M.C., Edmonson, M.N., et al. (2018). Genetic risk for subsequent neoplasms among long-term survivors of childhood cancer. J. Clin. Oncol. 36, 2078–2087.

46. Robison, L.L., Mertens, A.C., Boice, J.D., Breslow, N.E., Donaldson, S.S., Green, D.M., Li, F.P., Meadows, A.T., Mulvihill, J.J., Neglia, J.P., et al. (2002). Study design and cohort characteristics of the Childhood Cancer Survivor Study: A multi-institutional collaborative project. Med. Pediatr. Oncol. 38, 229–239.

47. Mailman, M.D., Feolo, M., Jin, Y., Kimura, M., Tryka, K., Bagoutdinov, R., Hao, L., Kiang, A., Paschall, J., Phan, L., et al. (2007). The NCBI dbGaP database of genotypes and phenotypes. Nat. Genet. 39, 1181–1186.

48. Crompton, B.D., Stewart, C., Taylor-Weiner, A., Alexe, G., Kurek, K.C., Calicchio, M.L., Kiezun, A., Carter, S.L., Shukla, S.A., Mehta, S.S., et al. (2014). The Genomic Landscape of Pediatric Ewing Sarcoma. Cancer Discov. 4, 1326LP–1341.

49. Perry, J.A., Kiezun, A., Tonzi, P., Van Allen, E.M., Carter, S.L., Baca, S.C., Cowley, G.S., Bhatt, A.S., Rheinbay, E., Pedamallu, C.S., et al. (2014). Complementary genomic approaches highlight the PI3K/mTOR pathway as a common vulnerability in osteosarcoma. Proc. Natl. Acad. Sci. 201419260.

50. Zhang, J., Bajari, R., Andric, D., Gerthoffert, F., Lepsa, A., Nahal-Bose, H., Stein, L.D., and Ferretti, V. (2019). The International Cancer Genome Consortium Data Portal. Nat. Biotechnol. 37, 367–369.

51. Heath, A.P., Taylor, D.M., Zhu, Y., Raman, P., Lilly, J., Storm, P., Waanders, A.J., Ferretti, V., Yung, C., Mattioni, M., et al. (2019). Abstract 2464: Gabriella Miller Kids First Data Resource Center: Harmonizing clinical and genomic data to support childhood cancer and structural birth defect research. Cancer Res. 79, 2464LP–2464.

52. Auton, A., Abecasis, G.R., Altshuler, D.M., Durbin, R.M., Bentley, D.R., Chakravarti, A., Clark, A.G., Donnelly, P., Eichler, E.E., Flicek, P., et al. (2015). A global reference for human genetic variation. Nature 526, 68–74.

