## Supplemental Methods for "Germline predisposition to pediatric Ewing sarcoma is uniquely characterized by inherited pathogenic variants in DNA damage repair genes"

**Study participants**

*Discovery cohort* – A total of 1180 unselected samples from patients with various sarcoma diagnoses were evaluated in the discovery phase of the study. These samples came from four data sources: St. Jude Cloud (Pediatric Cancer Genome Project, St. Jude Lifetime, Genomes for Kids, and Childhood Cancer Survivor Study; n = 1033)^1–5^, dbGaP study “Genomic Sequencing of Ewing Sarcoma” (phs000804.v1.p1; n = 26)^6,7^, dbGaP study “Osteosarcoma Genomics” (phs000699.v1.p1, n = 58)^8^, and ICGC study “Bone Cancer – UK” (BOCA-UK, n = 63)^9^. All 1180 samples met quality control thresholds for sequencing coverage and expected variant counts. After removing samples from related/ identical individuals, as well as samples not matching to one of 5 major continental ancestries (European, African, Admixed American, East Asian, and South Asian), samples from 1138 unique individuals were included in the discovery cohort.

*Ewing sarcoma validation cohort* – A total of 435 samples from patients with Ewing sarcoma family tumors were evaluated in the validation phase of the study. These samples were from the Gabriella Miller Kids First “Ewing Sarcoma – Genetic Risk” study (phs001228.v1.p1)^10^. 433 samples met quality control thresholds for sequencing coverage and expected variant counts. After removing samples from individuals that were related/ identical to those from the discovery cohort, as well as samples not matching to one of 5 major continental ancestries (European, African, Admixed American, East Asian, and South Asian), samples from 425 unique individuals were included in the validation cohort.

*Ewing sarcoma trio cohort* – A total of 301 patients (a subset of the 433 samples meeting quality control thresholds above) with Ewing sarcoma family tumors from the Gabriella Miller Kids First “Ewing Sarcoma – Genetic Risk” study had corresponding germline sequencing available from both parents (602 parents). These formed 301 full parent-proband sequenced trios, which were utilized for the inheritance analyses in this study.

*Uniqueness of Ewing sarcoma samples* – Based on available documentation, 19 patients with Ewing sarcoma from the discovery cohort in this study were previously included in a study of germline predisposition in Ewing sarcoma, and analyzed using approaches distinct from this current study^11^. The remainder of patients with Ewing sarcoma in the discovery and validation cohorts have not been previously analyzed in the context of a case-control germline predisposition study to our knowledge. Moreover, we have ensured that all patients with Ewing sarcoma in the discovery and validation cohorts of this study are unique and unrelated (i.e., do not have kinship coefficient above 0.125).

**Control cohorts**

Whole-exome sequencing data from six control cohorts with 24128 unrelated individuals without known cancer were used for our analyses. Exomes for controls were sequenced as previously described, and identically processed and analyzed in the same way as cases for this study. Subsets of these individuals were derived to create control cohorts matching the ancestry compositions of cases in the discovery and validation phases of this study. Under the assumption that the ancestry of parents would closely match those of probands in the validation Ewing sarcoma cohort, the same control cohort was utilized for pathway-based enrichment analysis among parents. The control cohorts came from the following sources: Autism Sequencing Consortium (phs000298), Framingham Cohort (phs000007), Multi-Ethnic Study of Atherosclerosis (phs000209), Lung Cohort (phs000291), in-house collection of exomes from National Heart, Lung, and Blood Institute “Grand Opportunity” Exome Sequencing Project (NHLBI GO-ESP), and the 1000 Genomes Project^12^. All germline analysis methods were also performed on the control cohorts unless stated otherwise.

***Quality Control***

**Sequencing and data harmonization**

The sequencing and bioinformatics pipelines used to generate the raw germline genomic data for all samples used in the discovery and control cohorts have been described previously^1–10^. For the Ewing sarcoma validation and trio cohorts, germline DNA was isolated from saliva. PCR-free libraries were constructed and sequenced on Illumina HiSeq X with 150 bp paired-end reads, and mean insert size of 325 bp. After library construction, the library was quantified on the LabChip GX. Fully ligated samples with minimum yield of 20ng passed quality control. Quantitative PCR was used to confirm concentration and check for dimer contamination.

Raw sequencing data was downloaded to Terra (<https://firecloud.terra.bio/>), a collaborative cloud-computing platform utilized for genomic analyses, developed as part of the NCI Cloud Pilot program and supported by the Broad Institute^13^. All raw sequencing Binary Alignment Map (BAM) files were realigned to the human reference genome b37 using BWA (version 0.7.15)^14^. In order maximize the number of samples available for this study, a combination of whole-exome sequenced (WES) and whole-genome sequenced (WGS) samples were used (Tables S1 and S2). For WGS samples, samtools (version 1.8)^15^ was utilized to convert sequencing to WES equivalents (retaining reads for exonic regions), based on target intervals from the Agilent SureSelect Human All Exon v2.0 bait set (Agilent Technologies, USA)^16^.

**Sequencing coverage**

The sample-wide mean coverage was calculated using GATK’s (version 3.7) tool “DepthofCoverage”^17^, and 10X coverage over evaluated intervals was used as the lower threshold for acceptable coverage.

**Variant count**

The sample-wide variant counts were determined using bcftools (version 1.9). Based on prior distributions of expected variant counts, and accounting for variability across ancestries, samples with whole-exome variant counts < 20,000 and > 40,000 were excluded.

**Cohort callset curation**

We merged the high-quality variants from all samples passing quality control steps pertaining to sequencing coverage and variant count into one Variant Call Format (VCF) file using GATK’s (version 3.7) tool “CombineVariants”. The “vt” tool (version 3.13) was used on the cohort VCF file to decompose multiallelic variants, followed by normalization of variants^18^. We evaluated the distribution of InDel length and ratio of transition to transversions using bcftools (version 1.9) tools’ “stats” and “plot-vcfstats” (Figure S3)^15^.

**Relatedness analysis**

We performed a relatedness analysis on the cohort VCF file in two steps. In the first step, we implemented GENESIS’s (version 2.12.0) tool PC-AiR to perform a Principal Components Analysis using genome-wide SNP data for the detection of population structure in a sample^19^. We then used GENESIS’s tool PC-Relate implemented in hail (version 0.2) (https://github.com/hail-is/hail) to estimate kinship coefficients between pairs of samples^20^. We removed one sample out of each pair that had a kinship coefficient above 0.125. Related samples were removed from the cohort VCF using GATK’s (version 3.7) tool “SelectVariants”.

**Ancestry inference**

We carried out ancestry inference separately for the following case cohorts: the pan-sarcoma discovery cohort, the osteosarcoma subset of the discovery cohort, the Ewing sarcoma subset of the discovery cohort, the rhabdomyosarcoma subset of the discovery cohort, and the Ewing sarcoma validation cohort. To perform ancestry inference, we merged each case cohort’s VCF file with our control VCF file and the 1000 Genomes reference samples using GATK’s tool “CombineVariants”. Using Hail (version 0.2), we filtered out variants in our callset with an allele frequency below 1% and variants that had a Hardy-Weinberg equilibrium p value less than 1 x 10^-6^. Additionally, we used the “ld_prune” method to filter out variants with a Spearman correlation threshold greater than 0.1. The “hwe_normalized_pca” method was used to obtain the principal component analysis (PCA) eigenvalues and scores. We performed uniform manifold approximation and projection (UMAP) on our callset using the package “umap” (version 0.4.3)^21^. We then trained a random forest classifier, using the sklearn (version 0.20.0) package’s “RandomForestClassifier” method, on the first ten principal components (PCs) and UMAP values from the 1000 Genomes cohort where we had continental ancestry annotations. We used the trained random forest classifier to assign one of the five 1000 Genomes defined super populations (European, African, Admixed American, East Asian, and South Asian) to each sample in our case and control cohorts (Figures S4 and S6). We then split our dataset into each continental ancestry and re-ran PCA. We visualized the first and second PC for each continental ancestry and manually selected clustering samples. Using the optmatch (version 0.9-14) package’s “pairmatch” function, we selected control samples that were closest to our case samples using PC 1-10 to create control cohorts matching the ancestry compositions of cases.

**Differential coverage analysis**

Using a two-sided Fisher exact test, we compared the mean fraction covered by more than 15 reads for each gene interval between the discovery case cohort and control cohort; we repeated this comparison between the validation case cohort and control cohort (Figure S2).

***Germline variant characterization***

**Detection of germline variants**

We called germline variants with a deep learning method, DeepVariant, which has shown superior sensitivity and specificity compared with a joint genotyping-based approach (version 0.8.0)^22,23^. For DeepVariant, we used the parameter “--model_type=WES”, which is recommended for Illumina Whole Exome Sequencing data. We only kept variants that were annotated with “PASS” in the “FILTER” column which means the probability of the non-reference genotype call was higher than for the reference genotype call (<https://github.com/google/deepvariant/issues/278>).

**Variant annotation and pathogenicity evaluation**

The cohort VCF file was annotated using the Variant Effect Predictor (VEP) (version 92) with the publicly available GRCh37 cache file with a custom plug-in to include a recent ClinVar database release (accessed in December 2019)^24^. Based on ClinVar database and VEP consequence annotations, all detected germline variants in cancer predisposition genes were classified into five categories: benign, likely benign, variants of unknown significance, likely pathogenic, and pathogenic using the American College of Medical Genetics (ACMG) guidelines^25^. Only putative loss-of-function, pathogenic, and likely pathogenic variants were included in this study (collectively referred to as pathogenic variants). Pathogenic germline variants in genes with nominal enrichment signal (p < .05), as well as those in DNA damage repair genes, were validated by examining BAM files using the Integrative Genomics Viewer (IGV version 2.8.6), marked as “True Positive” or “False Positive” depending on the depth of sequencing, the number of alternative allele reads, the variant allelic fraction (VAF), and the presence of artifacts at or around the examined variant site^26,27^ (Tables S4, S5, and S6).

**Statistical analyses**

Two-sided Fisher’s exact tests were used to calculate the odds ratios and confidence intervals (using “minimum likelihood correction”) for the enrichment of germline pathogenic variants in each of the examined cancer predisposition genes. Consistent with established statistical methods for multi-stage association studies, we implemented a permissive discovery phase analysis in which genes with p < 0.05 were considered to harbor nominal enrichment signal. The false discovery rate (FDR) was calculated using the Benjamini-Hochberg procedure; FDR < .05 was used as the threshold for enrichment reaching significance after multiple hypothesis testing correction. Top candidate genes were evaluated in the validation cohort. Fisher’s exact tests were also used for pathway-based enrichment analyses and to examine the association between germline mutational status and clinical characteristics. Odds ratios and confidence intervals were capped at 1000 for data visualization and reporting. Pooled analysis for *FANCC* was conducted using the fixed effect model, assuming that true effect size would vary between the discovery and validation cohorts due to sampling. All analysis was done in R (version 3.6.1) using the RStudio GUI (version 1.2.5001), and the “exact2x2”, “stats”, and “meta” packages were utilized.

**References**

1. McLeod, C., Gout, A.M., Zhou, X., Thrasher, A., Rahbarinia, D., Brady, S.W., Macias, M., Birch, K., Finkelstein, D., Sunny, J., et al. (2021). St. Jude cloud: A pediatric cancer genomic data-sharing ecosystem. Cancer Discov. *11*, 1082–1099.

2. Chen, X., Stewart, E., Shelat, A.A., Qu, C., Bahrami, A., Hatley, M., Wu, G., Bradley, C., McEvoy, J., Pappo, A., et al. (2013). Targeting Oxidative Stress in Embryonal Rhabdomyosarcoma. Cancer Cell *24*, 710–724.

3. Downing, J.R., Wilson, R.K., Zhang, J., Mardis, E.R., Pui, C.H., Ding, L., Ley, T.J., and Evans, W.E. (2012). The pediatric cancer genome project. Nat. Genet. *44*, 619–622.

4. Wang, Z., Wilson, C.L., Easton, J., Thrasher, A., Mulder, H., Liu, Q., Hedges, D.J., Wang, S., Rusch, M.C., Edmonson, M.N., et al. (2018). Genetic risk for subsequent neoplasms among long-term survivors of childhood cancer. J. Clin. Oncol. *36*, 2078–2087.

5. Robison, L.L., Mertens, A.C., Boice, J.D., Breslow, N.E., Donaldson, S.S., Green, D.M., Li, F.P., Meadows, A.T., Mulvihill, J.J., Neglia, J.P., et al. (2002). Study design and cohort characteristics of the Childhood Cancer Survivor Study: A multi-institutional collaborative project. Med. Pediatr. Oncol. *38*, 229–239.

6. Mailman, M.D., Feolo, M., Jin, Y., Kimura, M., Tryka, K., Bagoutdinov, R., Hao, L., Kiang, A., Paschall, J., Phan, L., et al. (2007). The NCBI dbGaP database of genotypes and phenotypes. Nat. Genet. *39*, 1181–1186.

7. Crompton, B.D., Stewart, C., Taylor-Weiner, A., Alexe, G., Kurek, K.C., Calicchio, M.L., Kiezun, A., Carter, S.L., Shukla, S.A., Mehta, S.S., et al. (2014). The Genomic Landscape of Pediatric Ewing Sarcoma. Cancer Discov. *4*, 1326 LP – 1341.

8. Perry, J.A., Kiezun, A., Tonzi, P., Van Allen, E.M., Carter, S.L., Baca, S.C., Cowley, G.S., Bhatt, A.S., Rheinbay, E., Pedamallu, C.S., et al. (2014). Complementary genomic approaches highlight the PI3K/mTOR pathway as a common vulnerability in osteosarcoma. Proc. Natl. Acad. Sci. 201419260.

9. Zhang, J., Bajari, R., Andric, D., Gerthoffert, F., Lepsa, A., Nahal-Bose, H., Stein, L.D., and Ferretti, V. (2019). The International Cancer Genome Consortium Data Portal. Nat. Biotechnol. *37*, 367–369.

10. Heath, A.P., Taylor, D.M., Zhu, Y., Raman, P., Lilly, J., Storm, P., Waanders, A.J., Ferretti, V., Yung, C., Mattioni, M., et al. (2019). Abstract 2464: Gabriella Miller Kids First Data Resource Center: Harmonizing clinical and genomic data to support childhood cancer and structural birth defect research. Cancer Res. *79*, 2464 LP – 2464.

11. Brohl, A.S., Patidar, R., Turner, C.E., Wen, X., Song, Y.K., Wei, J.S., Calzone, K.A., and Khan, J. (2017). Frequent inactivating germline mutations in DNA repair genes in patients with Ewing sarcoma. Genet. Med. *19*, 955–958.

12. Auton, A., Abecasis, G.R., Altshuler, D.M., Durbin, R.M., Bentley, D.R., Chakravarti, A., Clark, A.G., Donnelly, P., Eichler, E.E., Flicek, P., et al. (2015). A global reference for human genetic variation. Nature *526*, 68–74.

13. Birger, C., Hanna, M., Salinas, E., Neff, J., Saksena, G., Livitz, D., Rosebrock, D., Stewart, C., Leshchiner, I., Baumann, A., et al. (2017). FireCloud, a scalable cloud-based platform for collaborative genome analysis: Strategies for reducing and controlling costs. BioRxiv 209494.

14. Li, H., and Durbin, R. (2009). Fast and accurate short read alignment with Burrows-Wheeler transform. Bioinformatics *25*, 1754–1760.

15. Li, H., Handsaker, B., Wysoker, A., Fennell, T., Ruan, J., Homer, N., Marth, G., Abecasis, G., and Durbin, R. (2009). The Sequence Alignment/Map format and SAMtools. Bioinformatics *25*, 2078–2079.

16. Fisher, S., Barry, A., Abreu, J., Minie, B., Nolan, J., Delorey, T.M., Young, G., Fennell, T.J., Allen, A., Ambrogio, L., et al. (2011). A scalable, fully automated process for construction of sequence-ready human exome targeted capture libraries. Genome Biol. *12*, 1–15.

17. Depristo, M.A., Banks, E., Poplin, R., Garimella, K. V., Maguire, J.R., Hartl, C., Philippakis, A.A., Del Angel, G., Rivas, M.A., Hanna, M., et al. (2011). A framework for variation discovery and genotyping using next-generation DNA sequencing data. Nat. Genet. *43*, 491–501.

18. Tan, A., Abecasis, G.R., and Kang, H.M. (2015). Unified representation of genetic variants. Bioinformatics *31*, 2202–2204.

19. Conomos, M.P., Miller, M.B., and Thornton, T.A. (2015). Robust inference of population structure for ancestry prediction and correction of stratification in the presence of relatedness. Genet. Epidemiol. *39*, 276–293.

20. Conomos, M.P., Reiner, A.P., Weir, B.S., and Thornton, T.A. (2016). Model-free Estimation of Recent Genetic Relatedness. Am. J. Hum. Genet. *98*, 127–148.

21. McInnes, L., Healy, J., and Melville, J. (2018). UMAP: Uniform Manifold Approximation and Projection for Dimension Reduction.

22. Poplin, R., Chang, P.C., Alexander, D., Schwartz, S., Colthurst, T., Ku, A., Newburger, D., Dijamco, J., Nguyen, N., Afshar, P.T., et al. (2018). A universal snp and small-indel variant caller using deep neural networks. Nat. Biotechnol. *36*, 983.

23. AlDubayan, S.H., Conway, J.R., Camp, S.Y., Witkowski, L., Kofman, E., Reardon, B., Han, S., Moore, N., Elmarakeby, H., Salari, K., et al. (2020). Detection of pathogenic variants with germline genetic testing using deep learning vs standard methods in patients with prostate cancer and melanoma. JAMA - J. Am. Med. Assoc. *324*, 1957–1969.

24. McLaren, W., Gil, L., Hunt, S.E., Riat, H.S., Ritchie, G.R.S., Thormann, A., Flicek, P., and Cunningham, F. (2016). The Ensembl Variant Effect Predictor. Genome Biol. *17*, 1–14.

25. Richards, S., Aziz, N., Bale, S., Bick, D., Das, S., Gastier-Foster, J., Grody, W.W., Hegde, M., Lyon, E., Spector, E., et al. (2015). Standards and guidelines for the interpretation of sequence variants: A joint consensus recommendation of the American College of Medical Genetics and Genomics and the Association for Molecular Pathology. Genet. Med. *17*, 405–424.

26. Robinson, J.T., Thorvaldsdóttir, H., Winckler, W., Guttman, M., Lander, E.S., Getz, G., and Mesirov, J.P. (2011). Integrative genomics viewer. Nat. Biotechnol. *29*, 24–26.

27. Robinson, J.T., Thorvaldsdóttir, H., Wenger, A.M., Zehir, A., and Mesirov, J.P. (2017). Variant review with the integrative genomics viewer. Cancer Res. *77*, e31–e34.
