## Supplemental Figures for "Germline predisposition to pediatric Ewing sarcoma is uniquely characterized by inherited pathogenic variants in DNA damage repair genes"

### Figure S1

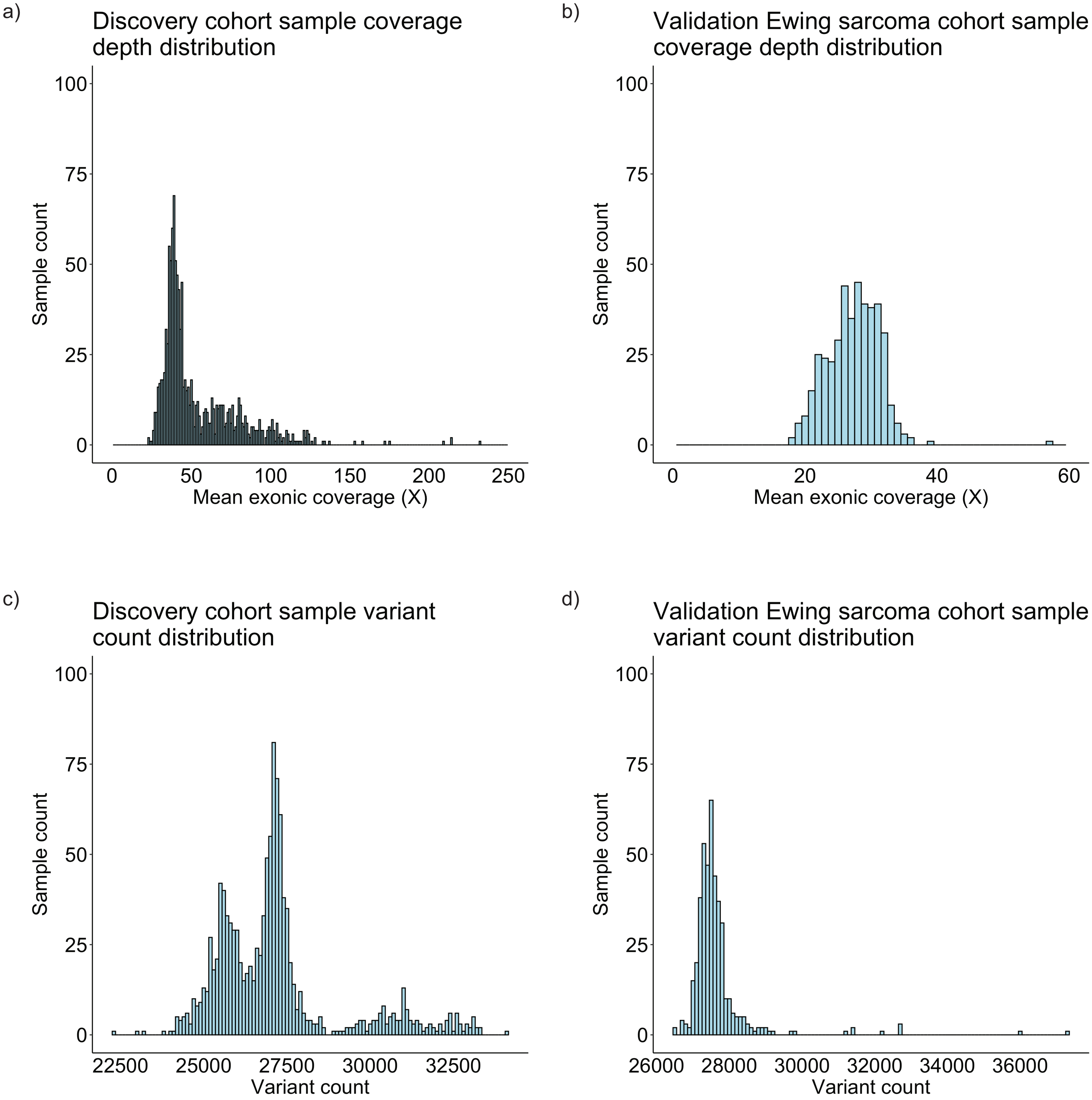


**Figure S1. Quality control: depth of coverage and sample-wide variant counts.** A, Distribution of mean exonic depth of coverage across discovery cohort. B, Distribution of mean exonic depth of coverage across Ewing sarcoma validation cohort. C, Distribution of total sample-wide variant count across discovery cohort. D, Distribution of total sample-wide variant count across Ewing sarcoma validation cohort.

### Figure S2

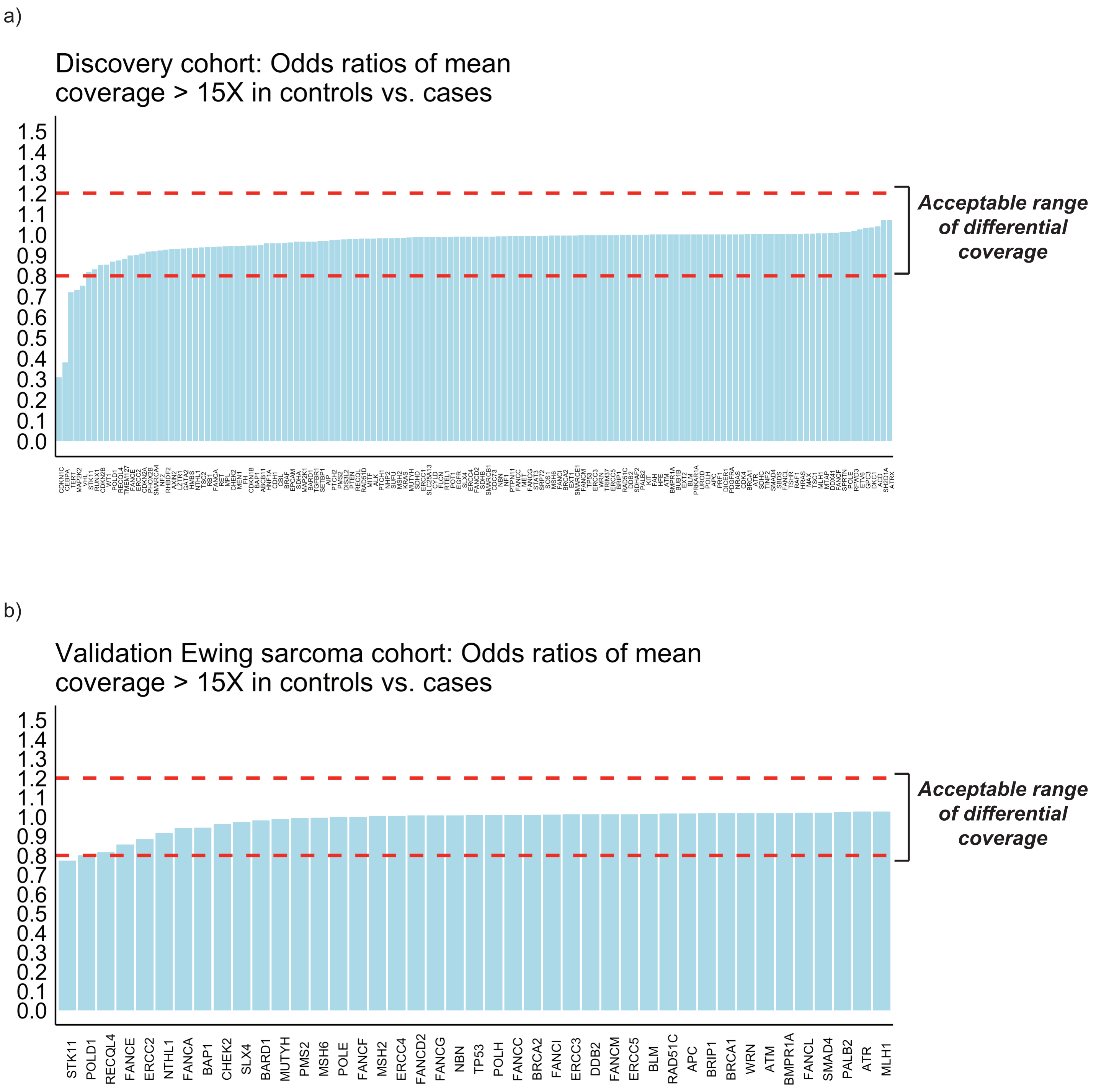


**Figure S2. Differential sequencing coverage of evaluated genes between controls and cases.** Differential coverage expressed as odds ratio of mean coverage over region of gene exceeding 15X in controls relative to cases. Odds ratios < 1 indicate greater coverage in cases relative to controls, whereas odds ratios > 1 indicate greater coverage in controls relative to cases. Odds ratios 0.8 - 1.2 used as range of acceptable differential coverage. A, Differential coverage across 141 genes evaluated in discovery cohort. B, Differential coverage across subset of 43 DNA damage repair genes evaluated in the Ewing sarcoma validation cohort.

### Figure S3

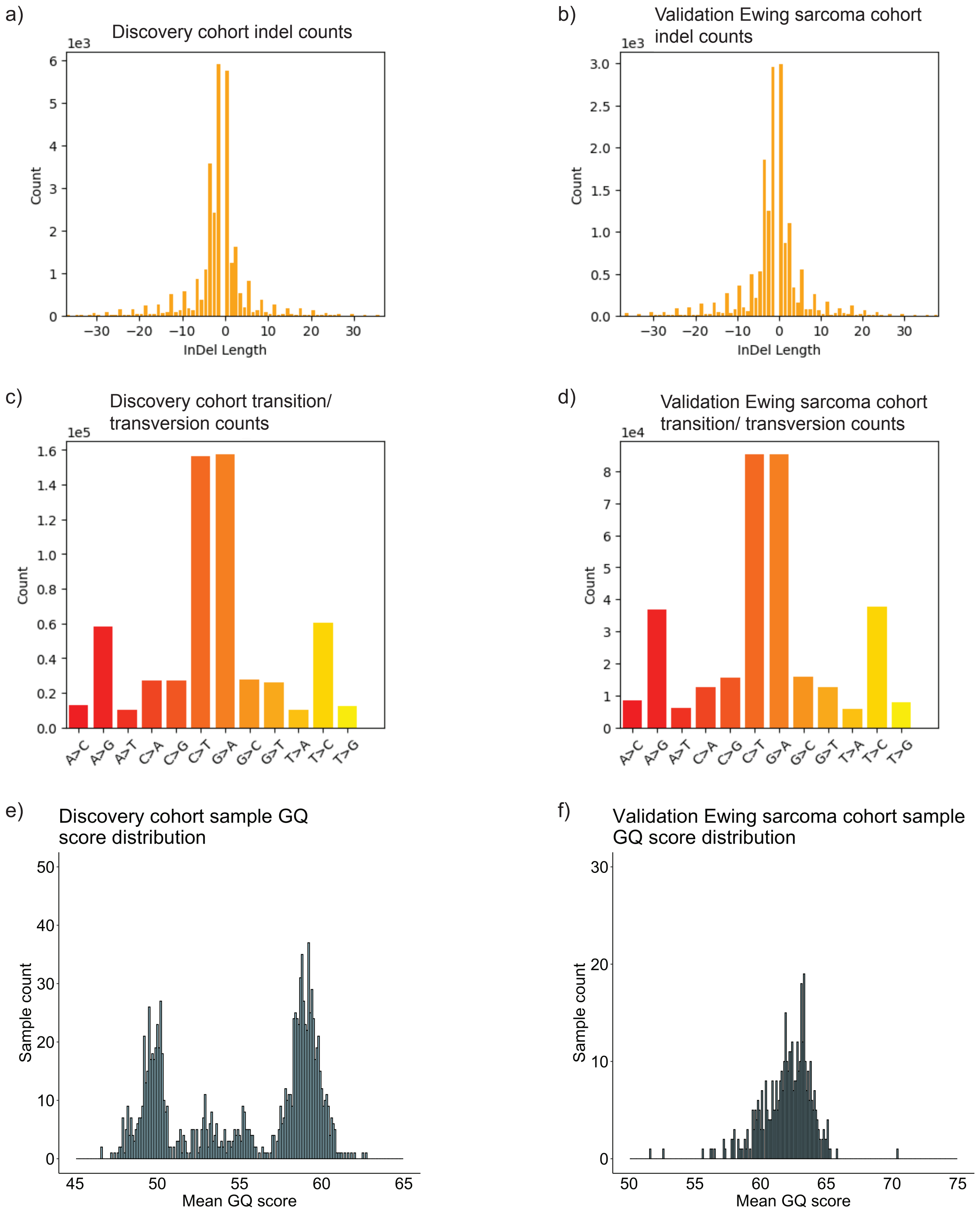


**Figure S3. Quality control: Indel counts, transition/ transversion counts, GQ score distribution.** Indel counts demonstrating relative counts for smaller indels in the A, discovery and B, Ewing sarcoma validation cohorts. Transition/ transversion counts illustrating transitions as being more common than transversion in C, discovery and D, Ewing sarcoma validation cohorts. Distribution of mean GQ scores (>30) for E, discovery and F, Ewing sarcoma validation cohorts, supporting high-confidence variant calls.

### Figure S4

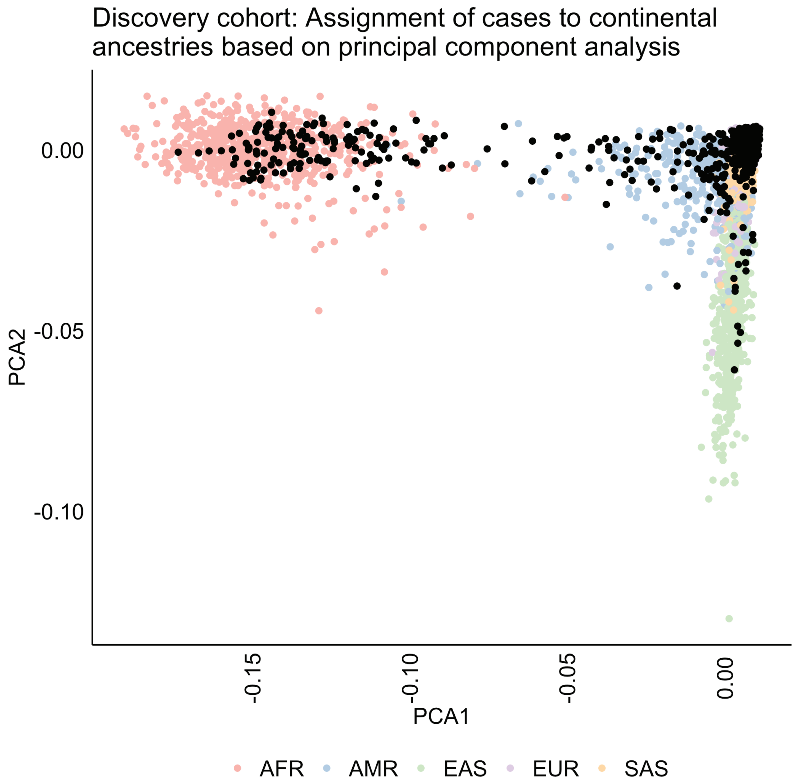


**Figure S4. Principal component analysis for continental ancestry assignment in the discovery cohort.** Visualization of the first two principal components. Control cohort is assigned to one of five major continental ancestries, indicated by the colored dots (AFR = African, AMR = Admixed American, EAS = East Asian, EUR = European, SAS = South Asian). Discovery case cohort is similarly assigned to one of five major continental ancestries (represented in black here for visualization).

### Figure S5

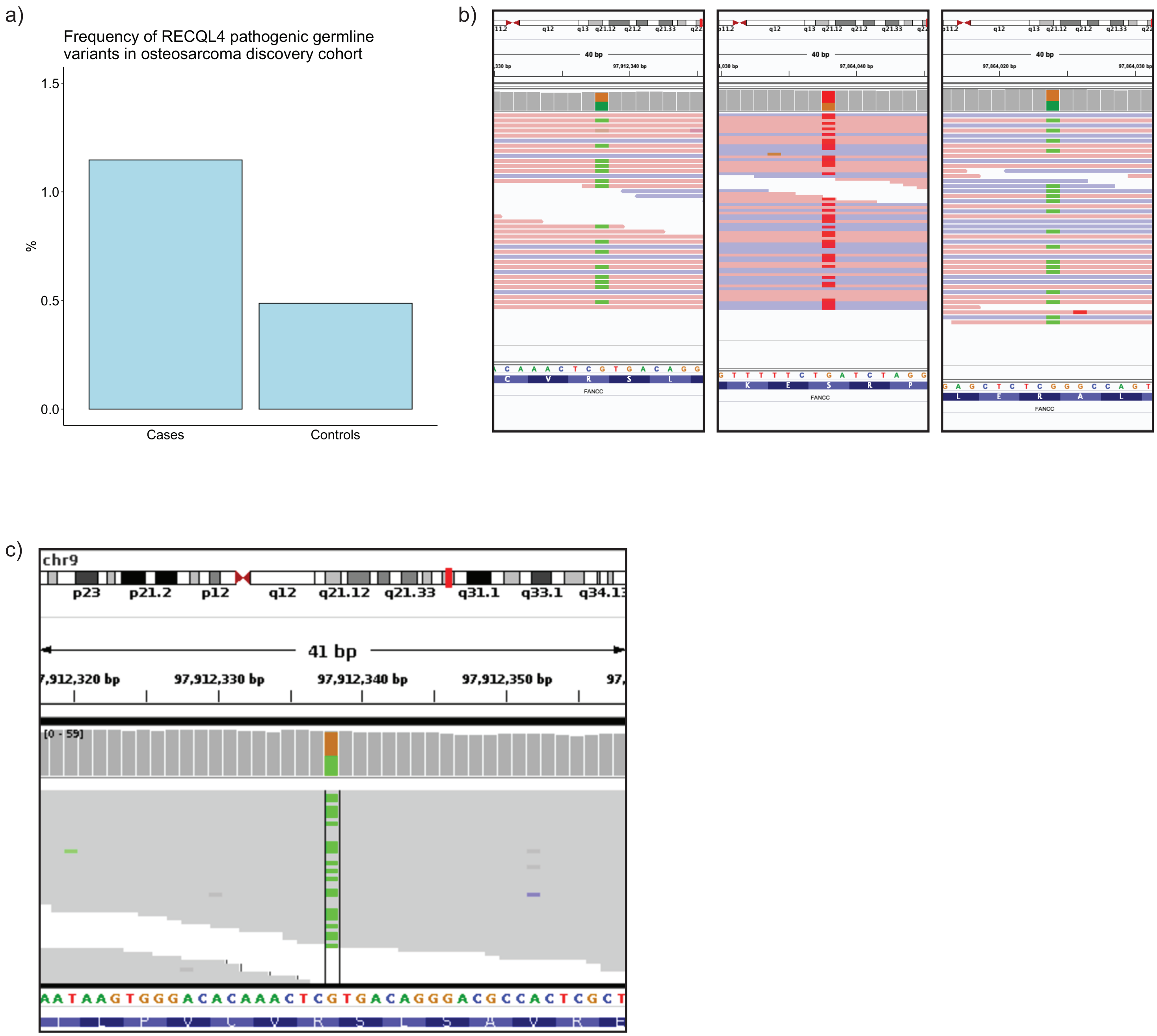


**Figure S5. Frequency and visualization of specific genes impacted by germline pathogenic variants in the discovery cohort.** A, Frequency of RECQL4 variants in the discovery osteosarcoma cohort is higher in cases relative to controls, but does not reach statistical significance. B, Visualization of germline pathogenic variants impacting FANCC in three different individuals with Ewing sarcoma in the discovery cohort. C, Matching tumor tissue from one of these individuals with Ewing sarcoma shows an identical heterozygous FANCC variant.

### Figure S6

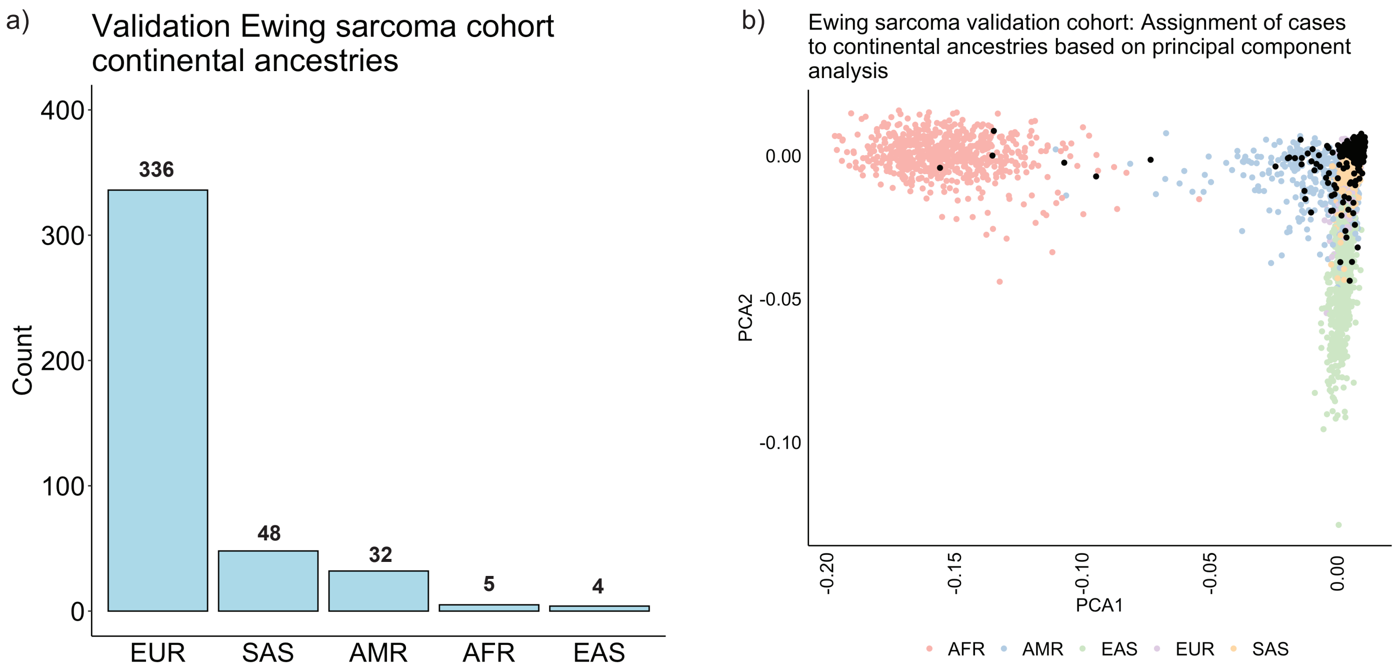


**Figure S6. Continental ancestry assignment by principal component analysis in the Ewing sarcoma cohort.** A, Ancestry composition of Ewing sarcoma validation cohort: 336 European cases (EUR), 48 South Asian cases (SAS), 32 Admixed American cases (AMR), 5 African cases (AFR), 4 East Asian cases (EAS). B, Visualization of the first two principal components. Control cohort is assigned to one of five major continental ancestries, indicated by the colored dots. The Ewing sarcoma validation case cohort is similarly assigned to one of five major continental ancestries (represented in black here for visualization).

### Figure S7

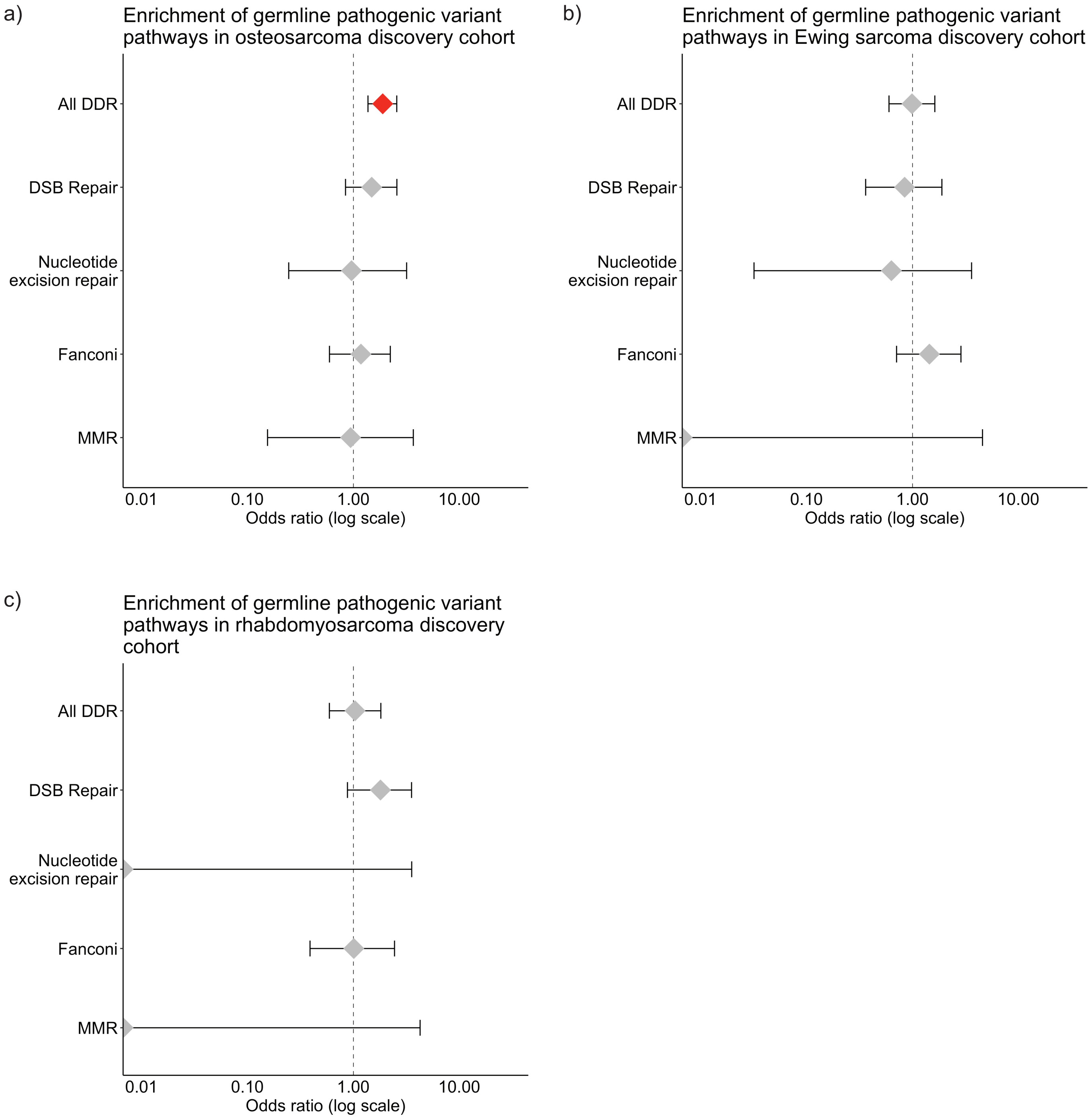


**Figure S7. Enrichment of pathogenic germline variants impacting DNA damage repair genes, analyzed in aggregate and by pathway.** Red: Significant at $p < 0.05$; Gray: Not significant. A, Pathogenic germline variants in DDR genes are enriched in aggregate (OR 1.9, 95% CI 1.4 - 2.6, $p < 0.05$) in the osteosarcoma discovery cohort, although specific pathways are not enriched. B, Neither DDR genes in aggregate or specific pathways are enriched in the Ewing sarcoma discovery cohort, possibly due to underpowering. C, Neither DDR genes in aggregate or specific pathways are enriched in the rhabdomyosarcoma discovery cohort, possibly due to underpowering.

### Figure S8

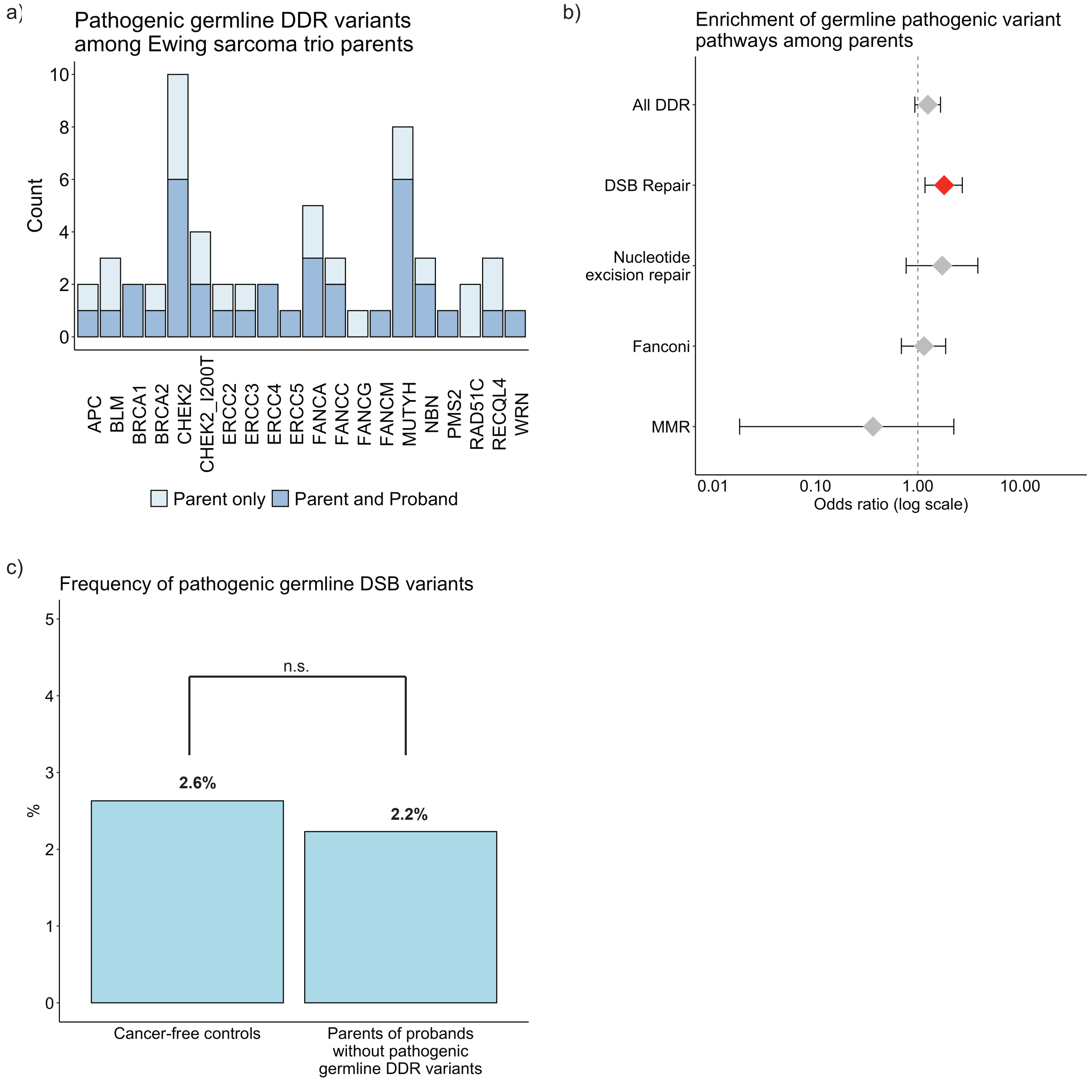


**Figure S8. Pathogenic germline variants in DNA damage repair genes among parents of probands with Ewing sarcoma.** A, 58 pathogenic germline variants in DDR genes among 602 parents of probands with Ewing sarcoma, stratified by concurrent presence or absence in proband. B, Pathogenic germline variants in double-strand break repair genes are enriched in parents of probands with Ewing sarcoma relative to controls (OR 1.8, 95% CI 1.2 - 2.7, $p < .05$). C, Frequency of germline pathogenic variants in DSB genes among parents of probands with Ewing sarcoma without identified germline DDR variants is comparable to the population frequency.

### Figure S9

a)

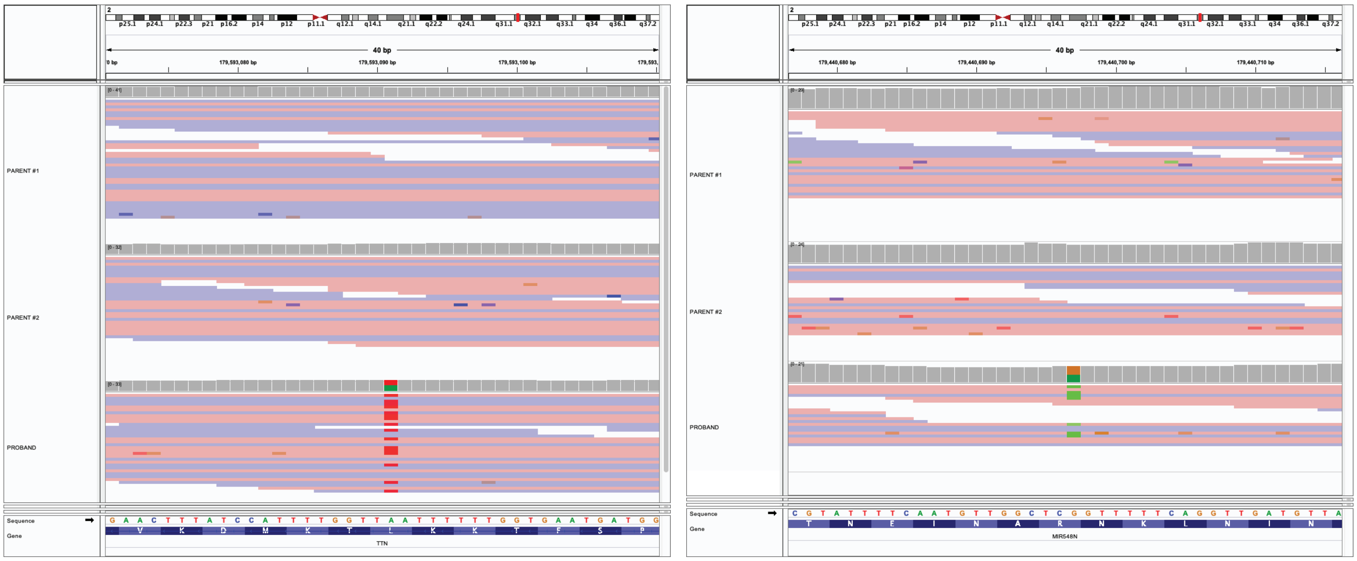


b)

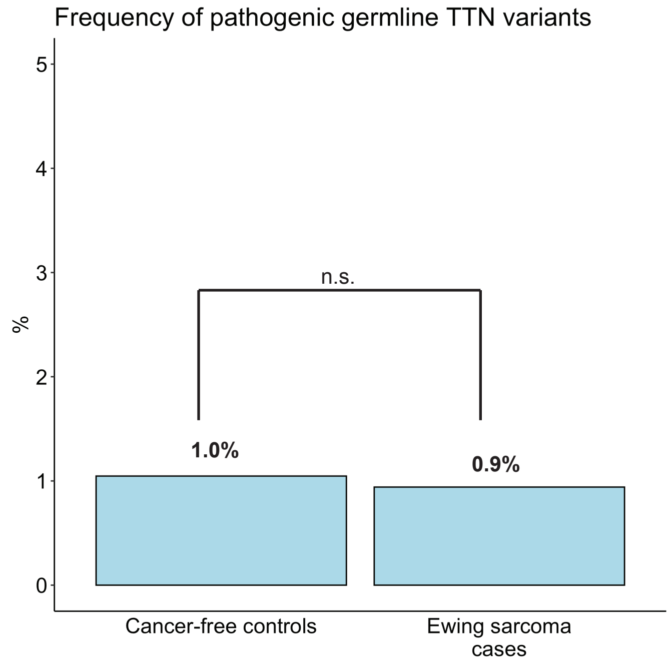


**Figure S9. Pathogenic germline variants in TTN.** A, While TTN is identified as being impacted by pathogenic de novo variants in 2 of 301 probands with Ewing sarcoma B, there is no overall enrichment in cases relative to controls to support its relevance to Ewing sarcoma pathogenesis.
